# Artificial Intelligence Agents in Mental Health: A Systematic Review and Meta Analysis

**DOI:** 10.64898/2026.04.21.26351365

**Authors:** Lexuan Zhu, Wenkong Wang, Zhiying Liang, Wenjia Tan, Bingyi Chen, Xinxin Lin, Zhengdong Wu, Huizi Yu, Xiang Li, Jiyuan Jiao, Sijia He, Guangxin Dai, Jiahui Niu, Yi Zhong, Yongbo Zheng, Jie Sun, Andi Han, Lingyao Li, Jiayan Zhou, Wenyue Hua, Ngan Yin Chan, Lin Lu, Yun Kwok Wing, Xin Ma, Lizhou Fan

## Abstract

The rapid rise of large language models (LLMs) and foundation models has accelerated efforts to build artificial intelligence (AI) agents for mental health assessment, triage, psychotherapy support and clinical decision assistance. Yet a gap persists between healthcare and AI-focused work: while both communities use the language of “agents,” clinical research largely describes monolithic chatbots, whereas AI studies emphasize agentic properties such as autonomous planning, multiagent coordination, tool and database use and integration with multimodal mental health data streams.

In this Review, we conduct a systematic analysis of mental health AI agent systems from 2023 to 2025 using a six-dimensional audit framework: (i) system type (base model lineage, interface modality and workflow composition, from rule-based tools to role-aware multi-agent foundation-model systems), (ii) data scope (modalities and provenance, from elicited self-report and chatbot dialogues to electronic health records, biosensing and synthetic corpora), (iii) mental health focus (mapped to ICD-11 diagnostic groupings), (iv) demographics (age strata, geography and sex representation), (v) downstream tasks (screening/triage, clinical decision support, therapeutic interventions, documentation, ethical–legal support and education/simulation) and (vi) evaluation types (automated metrics, language quality benchmarks, safety stress tests, expert review and clinician or patient involvement).

Across this corpus, we find that most systems (1) concentrate on depression, anxiety and suicidality, with sparse coverage of severe mental illness, neurocognitive disorders, substance use and complex comorbidity; (2) rely heavily on text-based self-report rather than clinically verified longitudinal data or genuinely multimodal inputs; (3) are implemented as single-agent chatbots powered by general-purpose LLMs rather than role-structured, workflow-integrated pipelines; and (4) are evaluated primarily via offline metrics or vignette-based scenarios, with few prospective, clinician- or patient-in-the-loop studies. At the same time, an emerging class of agentic systems assigns foundation models explicit roles as planners, retrieval agents, safety auditors or supervisors coordinating other models and tools. These multiagent, tool-augmented workflows promise personalization, safety monitoring and greater transparency, but they also introduce new risks around reliability, bias amplification, privacy, regulatory accountability and the blurring of clinical versus non-clinical roles.

We conclude by outlining priorities for the next generation of mental health AI agents: clinically grounded, role-aware multi-agent architectures; transparent and privacy-preserving use of clinical and elicited data; demographic and cultural broadening beyond predominantly Western adult samples; and evaluation pipelines that progress from offline benchmarks to longitudinal, real-world studies with routine safety auditing and clear governance of responsibilities between agents and human clinicians.

## Introduction

Mental health disorders affect more than one billion people worldwide [1], yet health systems remain chronically underresourced and have not adequately responded to population needs [2]. Mental health conditions are also highly heterogeneous: depressive and anxiety disorders, psychotic disorders, neurodevelopmental conditions and neurocognitive disorders differ markedly in symptomatology, course, comorbidity patterns and response to treatment. This heterogeneity implies that mental health care must be diversified and context-sensitive rather than one-size-fits-all. Combined with global shortages of specialist clinicians and barriers to accessing care, these features create a pressing demand for scalable, tailored mental health support and for tools that can augment rather than replace clinicians [1].

Recent advances in artificial intelligence (AI) make this demand particularly timely to examine. Since 2023, large language models (LLMs) have enabled conversational systems that can understand and generate natural language with high fluency across a wide range of topics [3], [4]. In parallel, AI systems have become increasingly *agentic* [5], moving beyond static chatbots to architectures that can plan multi-step actions, retrieve and synthesize external information, call tools and application programming interfaces (APIs), maintain memory across turns and coordinate multiple specialized components. Both non-LLM and LLM-based approaches can operate over multimodal inputs such as speech, images and structured clinical records. As a result, a rapidly growing body of work has begun to deploy AI agents for mental health screening, crisis support, longitudinal risk monitoring and conversational interventions in real-world or simulated settings [6]–[9]. At the same time, emerging analyses highlight serious concerns about safety, bias, privacy, accountability and alignment with clinical standards when such systems are used in high-risk contexts such as suicide prevention and automated assessment [8].

In this Review, we focus on *agentic AI systems for mental health*: systems in which one or more AI components autonomously select actions over a sequence of steps to advance mental health relevant goals. This broad definition includes (i) LLM-driven conversational agents that adapt over the course of a dialogue, (ii) multi-agent systems in which distinct agents debate, critique or supervise one another, and (iii) non-chat agentic components that operate in the background, for example, triage routers, safety sentinels or documentation assistants. By contrast, we do not treat purely static models that only produce one-shot predictions without any autonomous decision-making, such as a classifier that assigns risk scores to clinical notes but does not interact with users or tools, as “agents” in our framework. Clarifying this boundary is important because the design choices that make a system more agentic, for example, tool use, long-term memory or autonomous escalation, also introduce new clinical risks and governance requirements.

Despite intense interest and a surge of recent publications, the literature on AI agents for mental health is fragmented. Different communities use different terminology, for example, “chatbots”, “conversational agents”, “decision-support tools”, “multi-agent systems”, and evaluate systems under heterogeneous protocols, often with limited demographic or clinical reporting. Existing reviews [10]– typically focus on either pre-LLM digital interventions or on general-purpose LLM benchmarks, and therefore do not provide a consolidated picture of how modern agentic systems are actually designed, deployed and evaluated in mental health contexts. There is a need for a structured survey that connects technical architectures and training data to clinical targets, populations, downstream tasks and evaluation practices, and that makes explicit where current evidence is sufficient for deployment and where it remains preliminary or speculative.

To address this gap, we conduct a scoping review of recent AI agents for mental health, drawing on a curated set of studies identified through database and preprint server searches and systematically annotated using a shared schema. Building on this corpus, we propose a six-dimensional audit framework that characterizes each system along: (1) **System type**, for example, chat-based versus non-chat, single-agent versus multi-agent and their roles in a broader workflow, (2) **Data scope**: source, provenance and modality of data used for training and evaluation, (3) **Mental health focus**: target conditions or constructs, organized with reference to clinical taxonomies, (4) **Demographics**: age group, geography, sex and other reported characteristics of the populations involved, (5) **Downstream tasks**: including screening and triage, clinical decision support, therapeutic interventions, documentation generation, ethical and legal support, education and simulation, and evidence and benchmarking, and (6) **Evaluation types**: automated and human, including safety, usability and clinical validity. For each dimension, we summarize patterns across more than 300 recent papers, illustrate representative designs with concrete examples from the literature, and analyze how technical choices relate to clinical opportunities and risks.

Our contributions are threefold. First, we introduce a unified conceptualization of agentic AI systems for mental health that spans chat-based and non-chat settings and explicitly distinguishes agents from static predictive models. Second, we develop and apply a six-dimensional audit framework that organizes a diverse and rapidly expanding literature into a coherent landscape, revealing systematic skews, for example, towards depression and anxiety, adult populations and self-reported outcomes, and under-explored areas, such as severe mental illness, neurodevelopmental disorders and prospective clinical trials. Third, we critically examine current evaluation practices and governance approaches for mental health agents, highlighting misalignments between common technical metrics and clinically relevant endpoints, and outlining design and reporting recommendations for future work.

The remainder of this Review is organized as follows. We first introduce the six-dimensional audit framework and, in subsequent sections, examine each dimension in turn: system types and agentic roles, data scope and provenance, mental health focus and clinical settings, demographic coverage, downstream tasks and workflows, and evaluation types and evidence levels. Then, we discuss translational risks, regulatory and ethical considerations, and a research agenda for developing clinically responsible AI agents in mental health.

### System type: from rule-based tools to agentic foundation-model systems

In our framework, the *system type* dimension captures how mental health systems are instantiated in terms of their base models, user-facing interfaces and internal workflow composition. Concretely, this dimension answers three questions: (i) what learning paradigm and model family underpins the system (from rule-based to foundation models), (ii) how users and clinicians interact with the system (chatbot versus non-chat background agents), and (iii) how agentic roles are allocated and coordinated across components (non-agent pipelines, single-agent systems and multi-agent systems with different communication architectures). Together, these choices determine not only performance but also transparency, on-device feasibility, observability and clinical accountability.

### Base model lineage and clinical implications

We first organize systems by model lineage: rule-based, traditional machine learning, deep learning, small pre-trained models and general-purpose foundation models. Among studies published since 2023 in our annotated corpus, 238 implemented foundation models, 22 used small pre-trained models, 32 employed deep learning models, 13 used traditional machine learning models and 21 relied primarily on rule-based models. This distribution reflects a rapid shift towards foundation-model–based agents, but earlier paradigms remain important for interpretability, footprint-constrained deployment and well-controlled use cases.

#### Rule-based models

Recent reviews of mental health chatbots explicitly distinguish rule-based systems, traditional machine learning models, deep-learning models and large language model–based systems. Rule-based models generate responses from predefined rules using pattern-matching or finite-state logic [39]. For instance, one rule-based chatbot built on the Depression Anxiety and Stress Scale 21 (DASS-21) instruments achieved an average System Usability Scale (SUS) score of 77.2, suggesting that such structured chatbots can provide professionally acceptable and beneficial assistance to patients in constrained settings [40]. Similarly, another rule-based chatbot embedded core principles and techniques of cognitive behavioral therapy (CBT), enabling the system to deliver a structured, personalized adjunct to traditional therapy [41]. Clinically, rule-based systems offer high transparency and predictable behavior but limited adaptability outside their pre-specified rule sets.

#### Traditional machine learning models

As larger and more diverse datasets became available, investigators increasingly adopted traditional machine learning (ML) models to move beyond fixed rules. ML models learn decision boundaries directly from data instead of relying on hand-written rules [42]. Multiple studies use conventional classifiers such as support vector machines, random forests, decision trees and logistic regression to predict psychological health of pregnant women [43] and to detect stress and depression from behavioral or physiological signals [44]–[46]. These models can be comparatively data-efficient and interpretable, for example, through feature importance analysis but typically operate as single-task predictors without the broader agentic capabilities of modern systems.

#### Deep learning models

Building on this trend toward richer representations and better performance on unstructured data, deep learning models, particularly convolutional neural networks (CNNs) and recurrent neural networks (RNNs), have been widely adopted for mental health–related tasks [47]. CNNs have been used to analyze emotion from uploaded images within chat-based interventions [48], and deep models have been applied to refine replies in psychotherapeutic settings, achieving, for example, an average 47.6% reduction on an anxiety scale in one study [49]. RNN-based models such as long short-term memory (LSTM) networks have been integrated into frameworks like Rasa to enhance sentiment analysis capabilities of chatbots and support more coherent, context-aware conversations [50]. Reinforcement learning (RL), a core paradigm within deep learning, has been used both to shape agent behavior and to simulate psychopathology. Reinforcement learning from human feedback (RLHF) has been applied to therapeutic dialogue to encourage responses that prioritize empathy, moral considerations and human values [51], enabling more personalized interventions [52] and even tailored cognitive training, such as chess-based programs for cognitive impairment [53]. RL has also been used to simulate psychological disorders, including anxiety disorders, obsessive compulsive disorder [54] and cognitive impairment [53], providing controlled environments for probing symptom dynamics and potential treatment strategies.

#### Small pre-trained models

In parallel with higher-capacity architectures, small pre-trained models, for example, BERT- or RoBERTa-like encoders have been used both as end-user systems and as supporting components within larger pipelines. Some works use such models to generate empathetic responses or to classify user states in lightweight, privacy-sensitive deployments [55]. Others integrate small pre-trained models into LLM-based agents: for example, one 2024 study used a BERT model to annotate conversational text with emotions and guide speech synthesis, thereby enhancing the overall performance and expressiveness of an LLM-driven chatbot [56]. Clinically, these models can offer a pragmatic compromise between performance, interpretability and deployment constraints, for example, running on-device.

#### Foundation models

Foundation models, large models trained on massive corpora and adapted to diverse downstream tasks [57], now dominate recent mental health agent designs. PsychoLLM [58], for example, is a decoder-only model (Qwen1.5-14B-Chat) further trained via supervised fine-tuning (SFT) on multiple high-quality datasets including single-turn, multi-turn dialogues and knowledge-based question–answer pairs. Experiments show that PsychoLLM achieves strong performance in both English and Chinese on several proposed benchmarks, outperforming prior psychiatry-oriented LLMs such as EmoLLM [59], which is fine-tuned on LLaMA3-8B-Instruct, and MindChat, which is fine-tuned on Qwen-7B [60]. AutoCBT [61], fine-tuned on Qwen-2.5-72B for Chinese and Llama-3.1-70B for English, is reported to provide the best answer for more than 70% of bilingual questions in its evaluation. These systems illustrate how foundation models enable multi-lingual, multi-task mental health agents with rich reasoning capabilities, but they also raise new questions about data provenance, control, robustness and the clinical evidence required before deployment.

To rigorously compare these diverse lineages, we model the system selection as an optimization problem on a *Performance-Governance Plane*. Let each system type be characterized by its parameter count *P* and training data volume *D*. Following neural scaling laws, the clinical performance capability ℳ, such as reasoning depth or AUROC, scales logarithmically:

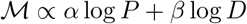

However, the deployment of such models incurs a “governance cost” 𝒞_*gov*_, which in a clinical mental health context is not merely a function of computational footprint, but of verification burden. We define 𝒞_*gov*_ as the divergence between the agent’s output distribution *P*_*θ*_(*y* | *x*) and the strict clinical gold standard *Q*_*clin*_(*y* | *x*), compounded by the cost of Human-in-the-Loop (*H*) intervention:

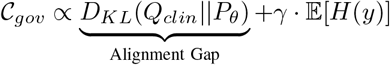

Here, *D*_*KL*_ denotes the Kullback-Leibler divergence—measuring the deviation of the model’s behavior from established protocols—while 𝔼[*H*(*y*)] represents the expected frequency of mandatory human verification, and *γ* is the cost of clinician time.

This stylized model captures the structural trade-off observed across our review. **Rule-based and ML models** naturally reside in a zone of minimal 𝒞_*gov*_ because their deterministic nature ensures *D*_*KL*_ ≈ 0 relative to the rules they encode, positioning them as the optimal choice for strictly regulated triage tasks despite their limited ℳ. By contrast, **foundation models** maximize ℳ through massive *P* and *D* but inherently introduce a non-zero Alignment Gap due to their stochasticity. This mathematical tension dictates the system’s realistic clinical role: as the base model’s capability increases, the governance cost 𝒞_*gov*_ rises, necessitating the layered safety overlays, such as critics, audit logs, and escalation gates, to artificially reduce the divergence from clinical standards.

### Interface modalities: chatbot front-ends versus non-chat background agents

The second aspect of system type concerns how users and clinicians interact with AI systems. We group interfaces into *chat-based* systems, which expose their capabilities through a conversational user interface, and *non-chat agents*, which operate primarily in the background or within existing clinical systems.

Across our corpus, nearly 91.74% of studies use chat interfaces. These systems leverage interactive dialogue to enhance accessibility, for example, by supporting multimodal inputs rather than restricting users to a single channel [19], [62]. One chatbot promoted diagnostic precision by integrating established assessment scales such as GAD-7, SRQ-20 and PHQ-9 directly into the conversational flow, dynamically selecting items based on user responses [48]. Other systems exploit the chat interface to capture additional signals beyond language, such as facial expressions, which are analyzed to infer mental health status [63], or to ingest electroencephalogram (EEG)–based emotion recognition for real-time affective monitoring [64]. From a clinical perspective, chat-based agents are well suited to psychoeducation, self-management support and adjunctive therapeutic interactions, but they also become the visible face of the system, concentrating both benefits and risks, for example, crisis responses, miscalibrated empathy.

Although chat remains dominant, non-chat systems also support clinically meaningful functions. Non-chat agents typically execute in the background: they monitor signals, retrieve evidence, generate documentation, manage scheduling or run safety checks, and then surface outputs within clinical systems rather than dedicated dialogue interfaces. For example, one system passively screened text-based media to detect users’ emotions without requiring active chat engagement [65], whereas another used audio data to estimate depression risk and provide risk scores in a clinical dashboard [66]. MindScope [67] further introduces a competitive debate mechanism inside a retrieval-augmented generation (RAG) pipeline and applies reinforcement learning to support decisions for detecting cognitive bias, demonstrating that substantial agentic reasoning can occur entirely behind the scenes. These designs clarify why *agents should not be conflated with chatbots*: many clinically important roles, such as triage routing, safety sentinels, documentation drafting or evidence retrieval, can be fulfilled by non-chat agents embedded into existing workflows. In summary, interface modality shapes who interacts with the system, patients, clinicians or both, at what points in the care pathway and with what expectations of control, transparency and support. A key implication for evaluation is that chat-based and non-chat agents require different usability and safety assessments, even when powered by similar underlying models.

### Workflow composition and agentic roles that determine reliability and accountability

To clarify the boundary between the diverse paradigms discussed in this review, we first formalize the mental health agent as a stateful decision process. While traditional classifiers establish a static mapping from input features to diagnostic labels (*y* = *f* (*x*)), agentic systems introduce dynamic state maintenance. Formally, an agent maintains a memory state *m*_*t*_ that evolves over time. At step *t*, given observation *o*_*t*_, the agent executes an action *a*_*t*_ ∼*π*_*θ*_(*a*_*t*_ *o*_*t*_, | *m*_*t*_).

Crucially, for clinical accountability, the state update cannot be a simple append operation. We refine the transition function *f*_*ϕ*_ to explicitly include a *Privacy Filter* (*F*_*priv*_) and a *Clinical Relevance Gate* (*G*_*clin*_):

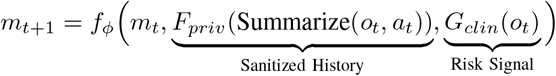

Here, *F*_*priv*_ decouples long-term clinical context from transient personally identifiable information (PII), while *G*_*clin*_(*o*_*t*_) ∈ [0, 1] is a scalar risk score that effectively locks the state into a safety protocol if a threshold is breached. This formulation mathematically distinguishes **non-agent pipelines** (where *m*_*t*_ is constant or absent) from **agentic systems** (where *m*_*t*_ accumulates context recursively via selective gating), foregrounding the reliability considerations that arise from path-dependent reasoning.

Building on this formalization, we consider how components are composed into workflows and which agentic roles they play. Concretely, we distinguish three levels of composition: *non-agent pipelines, single-agent systems* and *multiagent systems* (MAS) with flat, hierarchical, team or hybrid structures [68]. We also analyze which roles agents take on, such as personalization, risk assessment and tool use, and how these roles relate to reliability, observability and clinical accountability.

Most systems implicitly instantiate a variant of an *agent loop*: (i) sense and interpret input, for example, user text, speech or clinical context, (ii) retrieve relevant information or memory, (iii) plan next actions, (iv) execute actions, for example, generate responses, call tools, escalate to humans, and (v) reflect or update state. Non-agent pipelines implement only parts of this loop with fixed sequences of models, whereas single-agent and multi-agent systems allow more flexible control over planning, tool use and self-critique.

Non-agent systems typically comprise structured pipelines that perform prediction or classification tasks, for example, risk scoring or symptom detection without autonomous decision-making. Single-agent systems perform the entire loop within one model, often taking on a single agentic role such as “therapeutic counselor” or “documentation assistant”. By contrast, MAS allocate different roles to distinct agents or assign multiple agents to the same role to improve robustness. MAS can therefore accomplish complex target goals through coordinated interaction, often exceeding the capabilities of a single model operating alone [68]. Below, we highlight key roles and architectures.

#### Personalization

Personalization is one of the most common roles in mental health agents. Personalized user data provide a basis for LLM reasoning, supporting tailored diagnosis, monitoring and treatment. A multi-agent dual dialogue system [69], for example, reduces users’ cognitive load by analyzing conversations and offering personalized CBT exercises; the system is proposed as an augmented strategy for mental health providers. Many personalization approaches leverage persona construction or psychological scales. PsyDI [70] provides a personalized, in-depth chatbot for psychological measurement based on the Myers–Briggs Type Indicator (MBTI) framework. SocialSim [20] uses the Big Five model to infer help-seekers’ personality traits, constructs explicit seeker personas and generates empathetic, contextually appropriate responses tailored to those traits to reduce user’s psychological stress and provide concrete emotional support. Given the sensitivity of such data, privacy must be treated as a primary concern. SOULSPEAK [71] retrieves personalized context via a RAG framework while preserving user privacy through a dedicated module that uses named entity recognition to anonymize and later restore personal information, allowing the system to retain personalization benefits without exposing identifiers. Another study aiming to deconstruct depression stigma generates personalized causal knowledge graphs for individual users, enhancing intervention diversity and reducing unintended consequences [72].

#### Risk assessment

Risk assessment roles address both patient safety and system behavior. On the patient side, models are used to detect potential risks such as suicidal ideation from user text [73], [74]. On the system side, some work proposes single-agent evaluation frameworks to assess the safety and reliability of existing mental health chatbots, enabling post hoc calibration of deployed systems [75]. In addition, other studies have explored multi-agent safety evaluation frameworks, such as PsySafe [76], which assesses the safety of multiple interacting agents by quantifying risky behaviors and analyzing how their interactions may lead to unsafe outcomes. This line of research is increasingly important as more mental health systems adopt multi-agent architectures. Other approaches build single-agent refiners around LLM-generated answers, aiming to reduce overly cautious or exaggerated content while strengthening safeguards against harmful outputs [8]. Clinically, such roles are critical for escalation correctness, crisis management and reliable refusal behaviors.

#### Tool use and retrieval

Tool use in current systems is still dominated by relatively simple calls to LLM APIs [63], [77], domain-specific toolboxes (for example, a functional near-infrared spectroscopy analysis toolkit for anxiety and depression [78]) or AutoML systems such as TPOT for selecting appropriate downstream classifiers [66]. Richer tool infrastructures, such as ToolUniverse [79], remain relatively scarce in mental health. Some systems construct more structured tools: for example, one study built a dynamic diagnostic tree maintained by a dedicated tool agent that provides operational guidance to both doctor and patient agents [80]. Retrieval-augmented generation (RAG) plays an especially prominent role. In mental health applications, retrieval agents access specialized mental health knowledge bases [51], [81] and clinical practice guidelines [82]. Several studies construct novel datasets [22], [83], including psychotherapy dialogues, patient demographic profiles and personality trait data to improve RAG-based retrieval for specific tasks. Some MAS explicitly separate retrieval roles: for example, in a multi-agent dual dialogue system, dedicated retrieval agents inject external resources into the interaction before responses are finalized [69]. One study directly compares vector-based RAG with knowledge-graph–based RAG, showing that vector RAG offers dynamically updatable assistance, whereas graph-based RAG enhances retrieval robustness and supports complex, multi-turn interactions [84].

#### MAS architecture

Communication architectures determine how agents share information and coordinate.

To systematize the diverse multi-agent patterns reviewed, we model the system as a communication graph *G* = (*V, E*). Nodes *V* = {*A*_1_, …, *A*_*K*_} represent specialized agent policies (for example, therapist, critic), and edges *E* denote permissible interaction channels.

Crucially, we formalize the benefit of Multi-Agent Systems (MAS) not merely as task decomposition, but as **variance minimization**. Let distinct agents generate candidate responses with semantic variance *σ*^2^. The system reliability ℛ is inversely proportional to this variance:

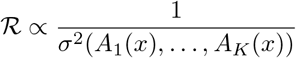

Consequently, the final clinical output *Y* is derived by an aggregation function Φ that acts as a conditional switch based on a safety threshold *ϵ*:

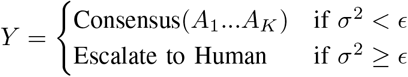

In this framework, **flat architectures** correspond to a fully connected graph where *E* supports peer-to-peer message passing, and Φ is typically a direct aggregation or turn-taking function. Flat architectures optimize for flow and empathy rather than variance control, operating without hierarchical oversight. They are commonly used in dialogue tasks where engagement takes precedence over strict clinical precision. CAMI [16] illustrates this for motivational interviewing: a client simulator and a counselor agent form a dyadic session to imitate real counselor behaviors. Similarly, SocialSim [20] uses a flat architecture for emotional support, where seeker and supporter agents take turns to enhance dialogue realism. While effective for simulation, the lack of a central Φ to compute *σ*^2^ limits their utility in high-stakes diagnostics.

**Hierarchical architectures** impose a directed tree topology where a root supervisor node (*v*_*sup*_) controls information flow. Here, Φ acts as a gated selection function: the supervisor explicitly evaluates the variance or quality of subordinate outputs before approval. ComPeer [85] adopts this structure, placing memory and scheduling blocks at the top level to coordinate dialogue generation, ensuring that responses remain contextually consistent (low *σ*^2^) over long interactions.

**Team architectures** operate on functional sub-graphs where peers share a specific goal. Here, Φ represents a complex *coordination protocol* (e.g., iterative debate or majority voting) rather than simple summation, maximizing robustness against individual errors. Mathematically, the objective of this protocol is to contract the semantic variance of the agents’ outputs through *N* rounds of interaction, such that 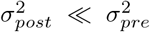. Team architectures involve collaboration among peer agents with specialized functions that share a common goal but operate without strict hierarchy. By integrating outputs through cooperative mechanisms such as debate, team architectures can enhance robustness and solution quality, reducing the risk that errors in a single “leader” propagate unchecked. MentalAgora [86] uses multi-agent debating for mental health support: agents assess user queries from different perspectives and a counselor LLM synthesizes the debate history and user data to produce personalized responses. MoodAngels [22] employs three diagnostic agents with varying levels of historical dependence, with Judge and Debate agents consolidating their outputs to obtain more accurate and robust psychiatric assessments (effectively minimizing *σ*^2^ via consensus).

**Hybrid architectures** utilize a dynamic topology *G*_*t*_. The system switches its structural configuration (e.g., from hierarchical planning to flat execution) based on the current interaction stage *t* or memory state *m*_*t*_, effectively optimizing the trade-off between control and creativity. This dynamic transition can be formalized as *G*_*t*+1_ = Ψ(*G*_*t*_, *m*_*t*_), where Ψ is a topology selection policy conditioned on clinical context. Hybrid architectures combine two or more patterns, enabling systems to switch structures across tasks or stages. ESC-Judge [23] adopts a hybrid architecture with three main stages: role construction, emotional-support conversation simulation and structured comparison, using hierarchical structures in earlier stages and a team-based architecture in the final comparison stage. AutoCBT [61] also uses a hybrid design: the overall system is hierarchical (user at the top, counselor in the middle, supervisors at the bottom), but the supervision component itself operates in a flat configuration, with supervisors providing parallel feedback on the counselor’s responses. MADP [87] maintains a hybrid multi-stage process: in Stage 1, dialogues are generated by multi-agents organized in a team architecture; in later stages, the system switches to a flat architecture for planning and response generation.

#### Representative systems with roles

MAGI [21] is a representative team-based MAS designed for psychiatric assessment interviews. It comprises four agents with distinct roles: a navigation agent controlling dialogue flow and output format, a question agent rephrasing questions to maintain engagement while preserving diagnostic intent, a judgment agent aligning free-form conversation with structured diagnostic criteria in real time, *effectively minimizing the Alignment Gap (D*_*KL*_*) by structured “sanitization”*, and a diagnosis agent synthesizing evidence into DSM-5–compliant diagnoses. Another representative MAS, MetaMind [88], aims to imitate human-like social reasoning. MetaMind deploys three agents: a theory-of-mind agent that generates hypotheses about the user’s mental state based on metacognitive theories, domain agents that integrate cultural norms and ethical constraints into these hypotheses, *functioning as variance constraints to ensure output safety (σ*^2^ < *ϵ)*, and a response agent that produces empathetic, coherent replies grounded in the refined mental-state models. Empirical results indicate that MetaMind achieves substantial improvements in real-world social scenarios and theory-of-mind reasoning performance.

Across these designs, workflow composition and agentic role allocation directly affect system reliability ℛ and accountability. Architectures that explicitly separate roles such as safety monitoring, retrieval and diagnosis can support clearer governance through *Local Variance Monitoring*, but they also introduce additional interaction failure modes (e.g., coordination errors in edge set *E*) that increases the global governance cost 𝒞_*gov*_ and must be carefully studied, particularly in high-stakes mental health applications.

### Technical realization and architectural spectrum

Tables I and II operationalize the theoretical framework through a technical audit of representative systems (2023–2025), decomposing them into their topology (*G*), base models (*θ*), and reasoning patterns (*π*). Analysis of these components reveals a marked trajectory toward **Specialized Role Distribution**: advanced systems like MoodAngels and ClinPreAI have transcended generic “assistant” personas in favor of discrete clinical functions, such as “Judge,” and “Police”, a specialization that necessitates complex debate or hierarchical topologies. Parallel to this structural shift is the adoption of **Reasoning-Driven Architectures**, where architectures such as AutoCBT and PsySafe replace single-turn generation with iterative “Draft-Refine” or “Self-Reflection” loops (*π*), deliberately trading inference latency for higher safety. Supporting these evolving capabilities is a transition to **Hybrid Memory Integration**, exemplified by MDD-5k, which augments simple context windows with structured Knowledge Graphs and Vector RAG to ground agent outputs in verifiable medical standards. Collectively, these engineering trends signal that the field is maturing from monolithic chatbots into modular, role-aware agentic ecosystems.

## Data scope: modalities and provenance that underpin clinical credibility

The *data scope* dimension characterizes what information mental health agents are trained and evaluated on, and how that information is obtained. We consider two axes: *modality*, for example, text, audio, image, video and physiological signals, and *provenance*, for example, clinically collected data, elicited evidence, interactive conversations and synthetic data. Together, these axes determine how well a system’s training and evaluation data support its claimed clinical use, including the realism of scenarios, the privacy risks involved, and the potential for bias or mismatch with target populations.

### Modalities: text-dominant systems and limited genuine multi-modality

Across our annotated corpus, the majority of systems remain effectively unimodal. One hundred eighty-six studies use exclusively text-based inputs, whereas only 26 explicitly incorporate multimodal data spanning combinations of text, images, audio and video. Moreover, even among studies that claim multimodal designs, many convert non-textual inputs, such as images, audio or video into text representations before processing, so that downstream reasoning is still performed by text-only models rather than by architectures with genuine cross-modal reasoning capabilities [96]–[98]. For example, several works transcribe speech into text for sentiment analysis or risk scoring instead of using acoustic prosody directly, and others use vision models only to generate textual labels that are then passed to LLMs.

A smaller set of systems implements *true* multimodal pipelines, where models jointly process audio, video and textual context. For instance, multimodal counseling agents combine text, audio and video streams to support psychological interventions and richer behavioral analysis at scale [56]. Other systems fuse text and facial-expression features to infer affective states [63], integrate electroencephalogram (EEG) signals with conversational cues for emotion recognition [64], or build depression-detection models that operate directly on audio recordings rather than transcriptions [66]. These designs illustrate the potential of multimodality to capture aspects of mental health, such as prosody, facial micro-expressions and physiological arousal, that are inaccessible from text alone, but they remain comparatively rare and are often evaluated on small or convenience samples.

From a clinical perspective, the current text-dominant land-scape has two implications. First, many agents making claims about nuanced affect or nonverbal behavior do so without systematically leveraging non-text signals. Second, systems that do incorporate richer modalities often face additional challenges in data collection, annotation and privacy, which limits sample sizes and diversity. As a result, there is a gap between the multimodal nature of real clinical encounters and the predominantly text-based data that underpin most mental health agents today.

### Data provenance and levels of evidence

For data provenance, we observe four main sources in our corpus:

1. **Clinically collected data**, such as electronic health records (EHRs), laboratory results and physiological signals.
2. **Elicited evidence**, including self-report questionnaires, structured assessments and standardized interviews, which can be situated within established evidence hierarchies.^1^
3. **Interactive conversation logs** from therapy, counseling or chatbot sessions.
4. **Generated or synthetic data**, such as LLM-simulated dialogues or synthetic patient trajectories.

Using our annotation schema, we find that more than one-third of recent systems rely on some form of generated or synthetic data, whereas a much smaller fraction use clinically collected EHRs or lab tests, and a substantial subset depend on elicited evidence such as screening scales and structured interviews. For example, several population-scale screening and monitoring studies use AI-enabled conversational agents to administer self-report instruments and collect high-volume questionnaire data for mental health risk assessment and progression modeling [6], [7]. Other work evaluates mental health chatbots using Rasch-based analyses of patient-reported outcomes, treating elicited questionnaire responses as the primary data source for assessing construct validity and response quality [99].

Each provenance category affords different strengths and limitations. Clinically collected data offer high ecological validity and alignment with real-world workflows but raise stringent privacy, governance and access challenges. Elicited evidence provides structured, interpretable measurements, for example, PHQ-9, GAD-7, DASS-21 but is often cross-sectional and may underrepresent marginalized groups. Conversation logs capture interactional dynamics over time but are expensive to curate and annotate at scale. Synthetic data can be produced in large quantities and used for stress-testing or pretraining, but their fidelity to real clinical populations depends on how well they are grounded in empirical evidence and expert oversight. For mental health applications, it is therefore crucial that claims about screening performance, decision support or therapeutic impact are commensurate with the provenance and evidentiary strength of the underlying data.

### Privacy, ethics and synthetic clinically grounded datasets

When creating datasets for mental health applications, privacy and ethics are central concerns. The most common strategy is to anonymize patient data before use [100], for example by removing direct identifiers and obfuscating free-text fields. However, several studies go further by synthesizing clinically grounded datasets from real cases, aiming to preserve clinical structure while reducing direct privacy risk.

MDD-5k, for instance, constructs a diagnostic conversation corpus from real, anonymized patient cases, diagnoses and outcomes, then synthesizes all concrete values into simulated diagnostic dialogues for downstream modeling [80]. Similarly, KMI simulates motivational interviewing by first training a model to imitate therapists’ behavioral choices and then using LLMs with prompt engineering to generate utterances, producing a synthetic, MI-grounded dataset of 1,000 high-quality Korean dialogues [93]. Other synthetic corpora use LLMs to simulate “artificial users” and counselors under controlled conditions, enabling systematic exploration of safety failures, empathy and adherence to therapeutic techniques without exposing real patients [87], [101].

These approaches illustrate a promising direction: using synthetic data to retain the clinical structure and decision logic of real encounters while mitigating re-identification risk. At the same time, they raise new questions about distributional shift and bias: synthetic dialogues may underrepresent edge cases, crisis scenarios or cultural nuances present in real populations. As with other provenance categories, the suitability of synthetic data depends on how systems are ultimately used, for example, as pretraining material, for stress-testing or as the sole basis for claims about clinical effectiveness.

Table III illustrates the strategic pivot toward agentic simulation to circumvent the “small data” bottleneck caused by medical privacy regulations. Moving beyond simple data augmentation, the field is defining a new standard through **Dual-Agent Simulation**, where systems like KMI and SocialSim orchestrate self-sustaining patient-therapist loops to autonomously produce high-volume corpora (e.g., 18k turns of MI dialogue). This generative capacity is increasingly paired with **Adversarial Safety Testing**, as seen in PsySafe’s use of “red-teaming” agents to simulate toxic behaviors for robustness checks. Crucially, to prevent these generative processes from drifting into hallucination, **Neuro-Symbolic Grounding** (exemplified by MDD-5k) anchors agent outputs to rigid diagnostic trees (e.g., SCID-5), ensuring clinical validity. Consequently, the field is moving toward a hybrid training paradigm where “agent-synthesized” curricula serve as the foundational bedrock before refinement on scarce human data.

**TABLE I.** Technical architecture and adaptation spectrum of representative mental health agent systems (2023–2025) – PartI.

| System | Clinical task | Topology ($G$) | Agent roles | Backbone and scale ($\theta$) | Adaptation | Memory and RAG ($m_t$) | Reasoning pattern ($\pi$) |
| --- | --- | --- | --- | --- | --- | --- | --- |
| **PsyDraw** [89] | HTP screening | Hierarchical (manager and specialists) | Manager; overall; house; tree; person | Claude-3.5; GPT-4o | Prompt engineering (manual) | Embedded in prompts | Task decomposition; two-stage (feature analysis, report) |
| **PsySafe** [76] | Safety defense (simulation) | Hierarchical; iterative loop | Doctor; police; contaminated patient | GPT-3.5; Llama-2 (7B, 13B, 70B) | Prompt engineering (dark traits injection) | Standard context window | Iterative optimization (doctor defense); self-reflection (police defense) |
| **MoodAngels** [22] | Mood disorder diagnosis | Team; debate | Angel.R; Angel.D; Angel.C; judge; debate agents | GPT-4o; DeepSeek-V3 | RAG-based ICL | Embedding: BGE-M3; source: DSM-5 + 2243 cases | Multi-round debate with granular analysis |
| **AutoCBT** [61] | CBT counseling | Dynamic routing (graph); star topology | Counselor (interface); supervisor agents (CBT experts) | Qwen-2.5 (72B); Llama-3.1 (70B) | Dynamic routing | Short-term and long-term memory (sliding window) | Draft-refine loop; CBT decomposition (5 standards) |
| **MentalAgora** [86] | Personalized counseling | Team; debate | Reframing counselor; positive regard counselor; solution-focused counselor | GPT-4; GPT-3.5; LLaMA-2 (13B) | Prompt engineering (role-play and persona) | Debate history buffer | Multi-round debate; attribute-weighted persona |
| **ClinPreAI** [36] | Postpartum depression risk | Iterative pipeline | Research; planner; coding; debugger; interpreter | Claude 4 Opus; Claude 4 Sonnet | Prompt engineering (LangGraph) | Internal KB (research module) | Iterative plan-code-debug loop with human feedback |
| **MentalRAG** [82] | Suicide risk and counseling | Hierarchical (parallel experts) | Conversation; anonymization; detection; report; proposal | Mental-LLaMA-3 (8B); GPT-4o; Mistral (7B) | QLoRA fine-tuning; RAG-based | Vector DB (ChromaDB); clinical guidelines (LLaMA-3 embeddings) | Parallel expert analysis with consensus aggregation |
| **MindEval** [38] | Psychotherapy simulation and benchmarking | Multi-agent sim (dyadic and evaluator) | Patient LM; clinician LM; judge LM | Patient: Claude 4.5 Haiku; judge: Claude 4.5 Sonnet; profile: GPT-5 | Prompt engineering (persona injection) | Full history and patient profile; few-shot ICL (judge) | Multi-turn interaction; automated evaluation |
| **MIND** [90] | Empathy training and cognitive restructuring | Hierarchical | Trigger; devil; guide; strategist | Gemini-2.0-flash | Prompt engineering (CoT) | Summary buffer | CoT; cyclic narrative control |
| **CAMI** [16] | MI | Single-agent (modular) | Counselor (state inference; topic; strategy; response) | GPT-4o; Llama-3.1 (70B) | Prompt engineering (zero- and few-shot) | Context window; static KB (topic tree) | STAR framework (state, topic, action, response) |

**TABLE II.** Technical architecture and adaptation spectrum of representative mental health agent systems (2023–2025) – Part II (Continued)

| System | Clinical task | Topology ($G$) | Agent roles | Backbone and scale ($\theta$) | Adaptation | Memory and RAG ($m_t$) | Reasoning pattern ($\pi$) |
| --- | --- | --- | --- | --- | --- | --- | --- |
| **PsychoGAT** [91] | Gamified psychometrics | Cyclical | Game designer; game controller; critic | GPT-4 | Prompt engineering | Recursive summarization | CoT; iterative critique-refine loop |
| **AMC** [92] | Depression and suicide risk diagnosis | Hierarchical; simulation | Psychiatrist; supervisor plugin; patient | GPT-3.5-turbo; GPT-4 (memory init) | Prompt engineering; RAG-based ICL (reflection) | Tertiary memory (history; EMR; skills); RAG: ICD-11; DSM-5-TR | Supervisor-guided instruction; reflection; majority voting ($k = 5$) |
| **MDD-5k** [80] | Synthetic diagnostic conversation | Hierarchical (tool agent manages) | Doctor; patient; tool agent | GPT-4o; Qwen2 (72B) | Prompt engineering (role-playing; persona) | Context buffer; KG (patient experience and symptom tree) | Neuro-symbolic dynamic diagnosis tree (hybrid rule, LLM) |
| **CA+** [17] | Long-term CBT and REBT | Hierarchical | Counselor (Albert Ellis); supervisor (evaluator) | GPT-4o | Prompt engineering | Structured DB (MySQL); RAG (Chinese textbooks) | Hierarchical recursive planning (goal-session-action) |
| **MAGI** [21] | Structured psychiatric interview (MINI) | Hierarchical; collaborative | Navigation; question; judgment; diagnosis | GPT-4o; Claude-3.5; GLM-Zero; DeepSeek-R1 | Prompt engineering (zero- and few-shot) | Dialogue history parsing; DSM-5 in system prompts | Finite state machine (interview tree); PsyCoT |
| **MADP** [87] | Mental health question answering (CBT-based) | Flat (collaborative team) | Explorer; empathizer; interpreter | Teacher: GPT-4o; student: LLaMA-3 (8B), GLM-4 (9B) | Prompt engineering (teacher); LoRA; instruction fine-tuning (student) | None | Multi-agent deductive planning (3-stage); reverse ABC analysis |
| **KMI** [93] | MI simulation | Flat (dyadic) | Therapist simulator; client simulator | GPT-4; T5-base (220M) | Prompt engineering (few-shot ICL); full SFT (forecaster) | Sliding window (6 turns) | Forecast-then-generate: MI forecaster, rule-based decision, generation |
| **Yang et al.** [94] | Construction safety and mental health | Flat (collaborative; debate) | Occupational safety and health specialist; Human resources advisor; worker peer | GPT-4o | RAG-based ICL; prompt engineering (two-tier chaining) | Shared history; RAG: FAISS and OpenAI embeddings (OSHA, labor docs) | Dynamic orchestration (sequential; parallel debate) |
| **Kampman et al.** [95] | Therapist support in text counseling | Flat | Propose response; recommend resources; analyze; summarize; empathetic rewrite | GPT-4o; Llama 3 (8B, 70B) | Prompt engineering; RAG | Full conversation context; RAG: FAISS (Mindline.sg content) | Human-in-the-loop orchestration; template-based prompting |

**Note:** $G$ : Agent Topology; $\theta$ : Base Model Parameters; $m_t$ : Memory State; $\pi$ : Reasoning Policy. **ICL**: In-Context Learning; **SFT**: Supervised Fine-Tuning; **RAG**: Retrieval-Augmented Generation; **CoT**: Chain-of-Thought; **PsyCoT**: Psychometric Chain-of-Thought; **KG**: Knowledge Graph; **EMR**: Electronic Medical Records; **HTP**: House-Tree-Person; **MI**: Motivational Interviewing; **REBT**: Rational Emotive Behavior Therapy; **MINI**: Mini-International Neuropsychiatric Interview.

**TABLE III.**
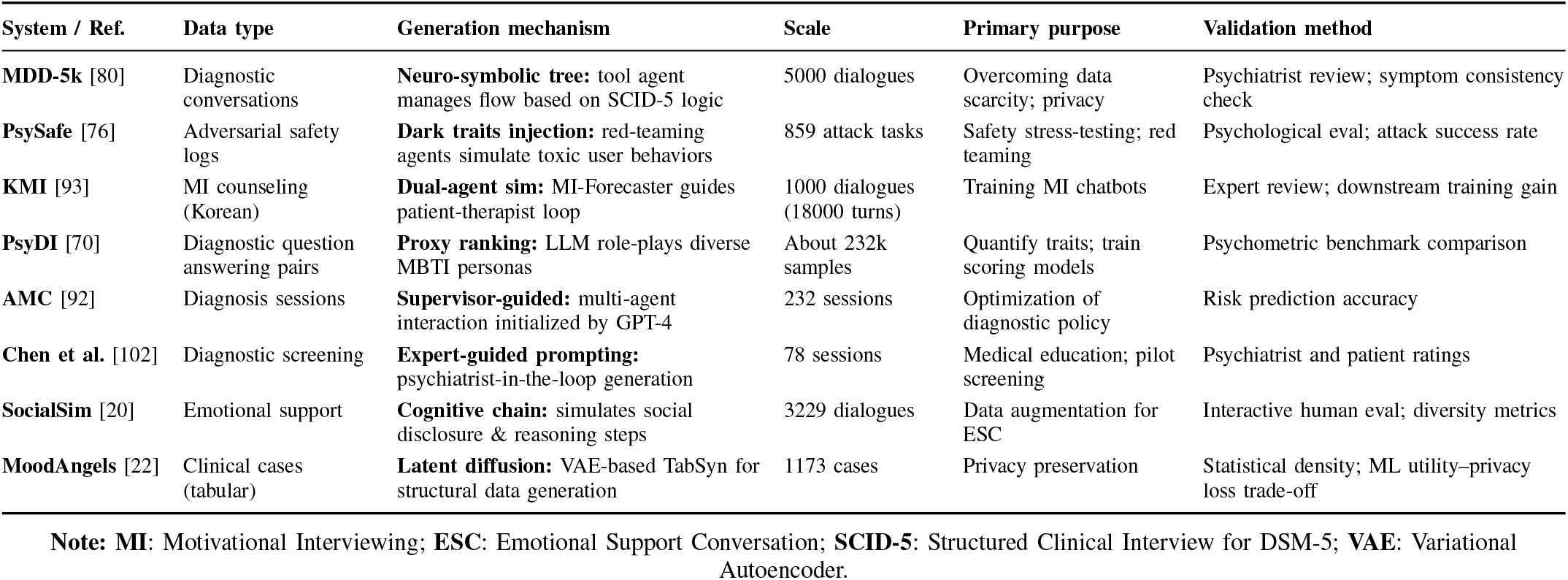
Landscape of synthetic data generation frameworks in mental health (2023–2025)

| System / Ref. | Data type | Generation mechanism | Scale | Primary purpose | Validation method |
| --- | --- | --- | --- | --- | --- |
| MDD-5k [80] | Diagnostic conversations | **Neuro-symbolic tree:** tool agent manages flow based on SCID-5 logic | 5000 dialogues | Overcoming data scarcity; privacy | Psychiatrist review; symptom consistency check |
| PsySafe [76] | Adversarial safety logs | **Dark traits injection:** red-teaming agents simulate toxic user behaviors | 859 attack tasks | Safety stress-testing; red teaming | Psychological eval; attack success rate |
| KMI [93] | MI counseling (Korean) | **Dual-agent sim:** MI-Forecaster guides patient-therapist loop | 1000 dialogues (18000 turns) | Training MI chatbots | Expert review; downstream training gain |
| PsyDI [70] | Diagnostic question answering pairs | **Proxy ranking:** LLM role-plays diverse MBTI personas | About 232k samples | Quantify traits; train scoring models | Psychometric benchmark comparison |
| AMC [92] | Diagnosis sessions | **Supervisor-guided:** multi-agent interaction initialized by GPT-4 | 232 sessions | Optimization of diagnostic policy | Risk prediction accuracy |
| Chen et al. [102] | Diagnostic screening | **Expert-guided prompting:** psychiatrist-in-the-loop generation | 78 sessions | Medical education; pilot screening | Psychiatrist and patient ratings |
| SocialSim [20] | Emotional support | **Cognitive chain:** simulates social disclosure & reasoning steps | 3229 dialogues | Data augmentation for ESC | Interactive human eval; diversity metrics |
| MoodAngels [22] | Clinical cases (tabular) | **Latent diffusion:** VAE-based TabSyn for structural data generation | 1173 cases | Privacy preservation | Statistical density; ML utility–privacy loss trade-off |

**Note:** MI: Motivational Interviewing; ESC: Emotional Support Conversation; SCID-5: Structured Clinical Interview for DSM-5; VAE: Variational Autoencoder.

### Clinically collected data and electronic health records

Clinically collected data, especially EHRs and physiological signals, provide a complementary route to realism. The incorporation of EHR-derived signals such as EEG, combined with multimodal data like interview audio, has been shown to improve mental health assessment by offering richer insight into real-time psychological and emotional states [64], [103]. However, unstructured elements of EHRs, particularly narrative clinical notes, can conceal valuable information and are difficult to use directly. This challenge has been addressed by methods that apply longitudinal patient-history summarization, multi-step reasoning and confidence-aware decision-making to uncover latent cognitive health signals and support detection of cognitive impairment [104].

EHR-linked physiological data also appear in phobia treatment studies using virtual reality (VR), where heart-rate measurements are captured in real time and used to calibrate anxiety levels during exposure exercises [105]. Other work combines wearable sensor streams with conversational data to build holistic monitoring systems for mood and anxiety, although such systems are still in early stages and often evaluated on small cohorts [106]. Overall, while clinically collected data offer the strongest alignment with real-world practice, they also demand the most stringent safeguards around consent, de-identification, governance and data linkage. For AI agents in mental health, using such data responsibly requires not only robust technical safeguards but also clear documentation of data provenance, cohort characteristics and limitations, so that downstream claims about screening, decision support or treatment impact are grounded in an appropriate level of clinical evidence.

## Mental health focus: ICD-11 mapping of topical coverage and blind spots

The *mental health focus* dimension captures which diagnostic groups and clinical constructs current systems target. We map reported conditions to ICD-11 groupings [107] to quantify topical skew and identify blind spots. Overall, a clear pattern emerges: many conversational-agent studies concentrate on broad “general psychiatry” use cases, typically positioning the system as a broadly accessible tool for emotional uplift, self-help or early screening. This means they focus on dealing with all mental health disorders within a single general-purpose system, rather than tailoring methods with regard to different disorder types.

A large subset of agents explicitly frame themselves as general-purpose mental health companions or peer-like supports. For example, one line of work incorporates an EmoCBT module that draws on core cognitive–behavioral therapy principles to deliver peer-like support for everyday mental well-being [108]. Other systems imitate human interaction through natural language processing and machine learning techniques to provide general psychoeducation and mood support [109]. JeevaAI further emphasizes a user-centered, inclusive and highly personalized approach designed to scale across diverse populations globally [110]. Moving beyond text-only interaction, Multimodal Emotion-Aware Conversational Agent (MEACA) extends general support by jointly modeling textual input, facial expressions and physiological signals to better infer users’ emotional states and respond more appropriately [111]. Similarly, Tessa combines LLM-based language understanding with an embodied conversational agent to incorporate both verbal and nonverbal cues, using CBT-informed guidance to provide general support across user needs [112]. These systems are typically labeled as “general psychiatry” in our annotation, and they dominate the corpus relative to disorder-specific agents.

When we restrict attention to studies that focus on specific mental health disorders (Figure 4), the literature is heavily skewed toward depressive disorders and anxiety disorders, with limited coverage of severe mental illness, neurocognitive disorders, substance use, sleep–wake disorders, neurodevelopmental conditions and complex comorbidity. In our coding, ICD-11 groups corresponding to depressive disorders and anxiety or fear-related disorders account for the majority of disorder-focused systems, whereas codes associated with psychotic disorders, bipolar disorders, neurocognitive disorders and substance use disorders appear only sporadically. This imbalance has direct implications for generalizability and safety: systems trained and evaluated primarily on depression and anxiety may behave unpredictably when used with populations experiencing psychosis, mania, cognitive impairment or substance misuse, even when marketed as “general-purpose” mental health agents.

**Fig. 1.**
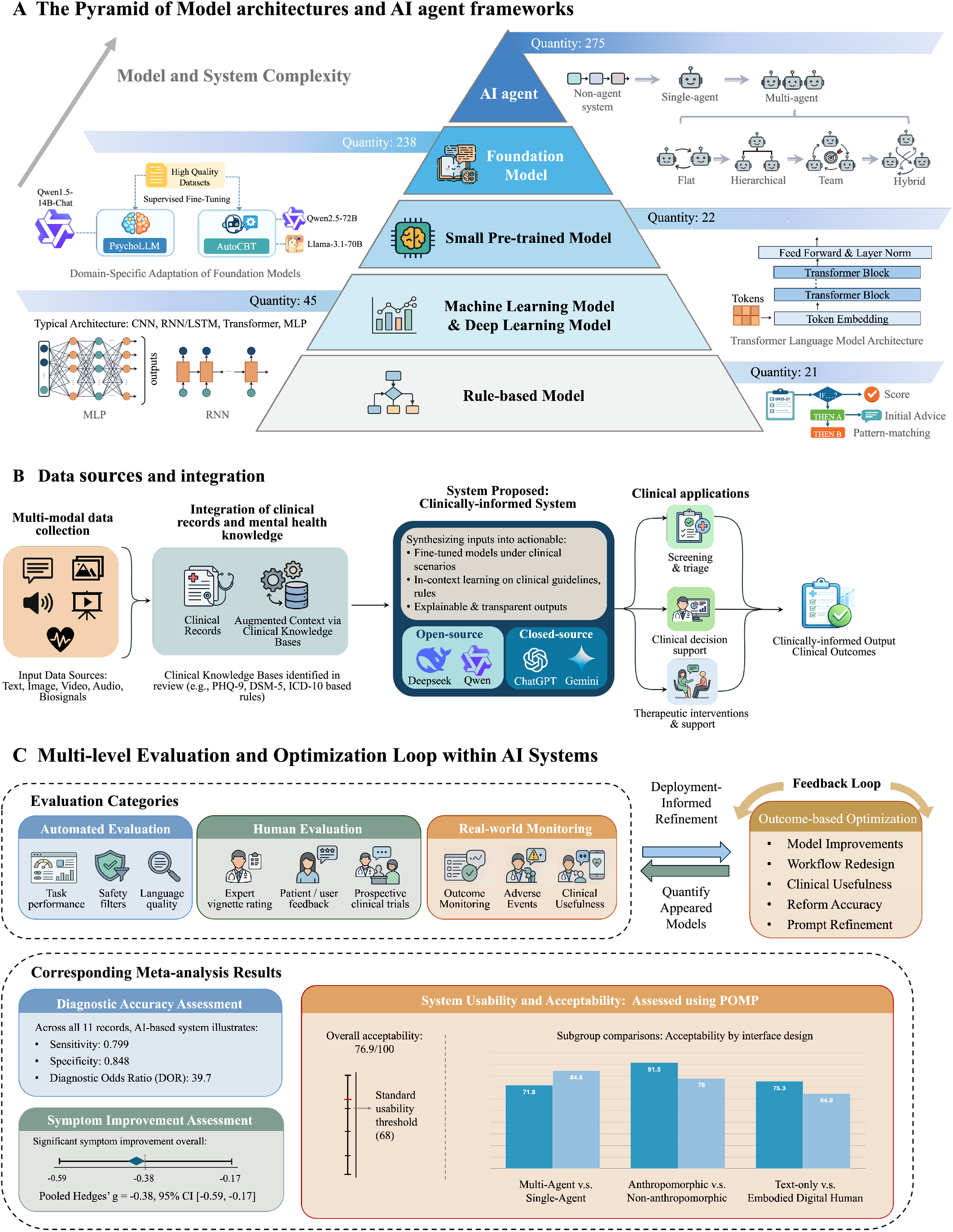
Landscape overview of AI agents in mental health. The taxonomy spans three dimensions: (1) The Pyramid of Model architectures and AI agent frameworks, detailing each categories by model and system complexity (from rule-based systems to AI agent), (2) Data source and integration, covering the workflow under the clinical setting from the inputs (text, image, video, audio and biosignals), to the integration of clinical records and knowledge, to the clinically-informed system, to clinical applications (downstream tasks) and finally the outputs; (3) Multi-level Evaluation and Optimization Loop within AI systems. Within the loop, three evaluation categories are specified (automated evaluation, human evaluation and real-world monitoring) with corresponding meta-analysis results. The mutual refinement and the quantification would enhance the performance of the proposed systems.

**Fig. 2.**
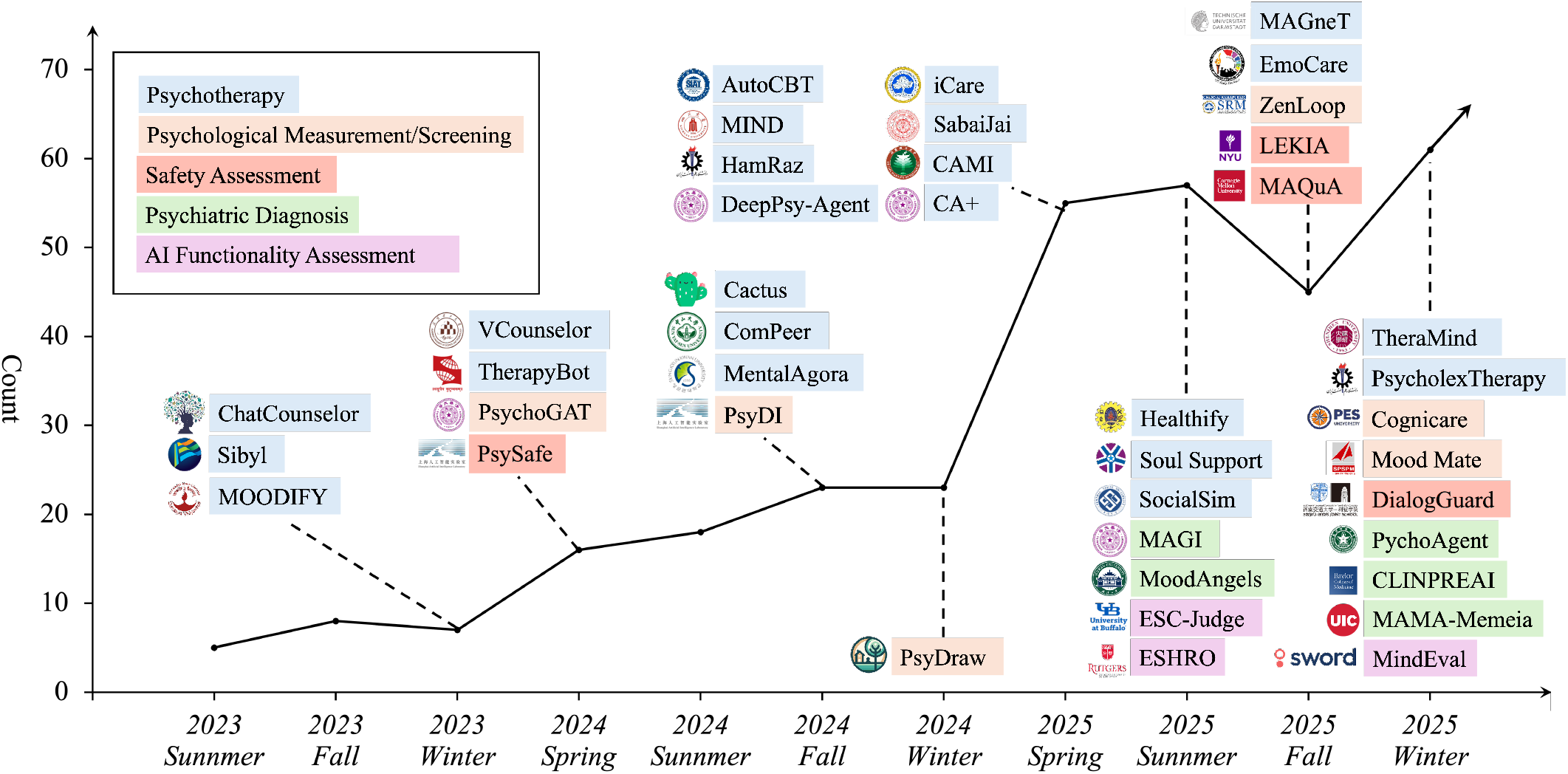
Growth of AI agent in mental health literature and representative exemplar systems, 2023–2025. The curve shows the cumulative number of papers over time (y-axis: paper count; x-axis: publication season by year), with dashed vertical lines marking calendar years. Colored labels highlight representative systems published in each period, grouped by their primary application domain: psychotherapy, psychological measurement/screening, safety assessment, psychiatric diagnosis and AI functionality assessment (legend, upper left).

**Fig. 3.**
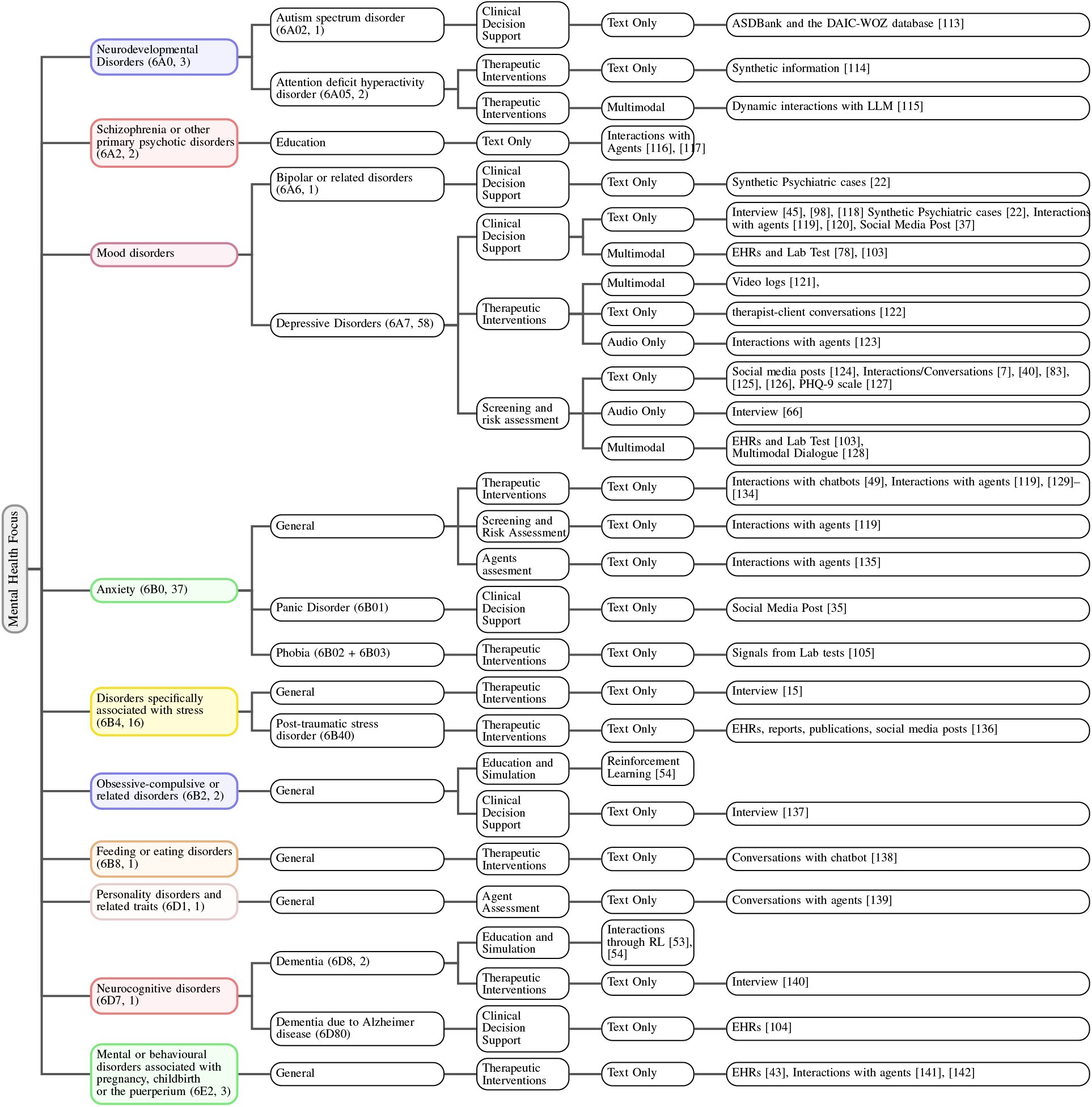
Hierarchical mapping of AI agents in mental health to ICD-11 diagnostic groups, downstream tasks, input modalities and evidence sources. High-level ICD-11 categories (for example, neurodevelopmental disorders, mood disorders, anxiety and stress-related disorders, neurocognitive disorders) are shown on the left, with disorder codes and the number of included papers. Branches expand to specific conditions (such as depressive disorders, panic disorder, post-traumatic stress disorder and dementia due to Alzheimer’s disease), associated task types (clinical decision support, therapeutic interventions, screening and risk assessment, agent assessment, education and simulation), dominant modalities (text-only, audio-only or multimodal) and representative data sources (for example, electronic health records, lab tests, social media posts, clinical interviews and interactions with agents or chatbots). Individual studies may appear on more than one branch when they target multiple disorders or task types.

**Fig. 4.**
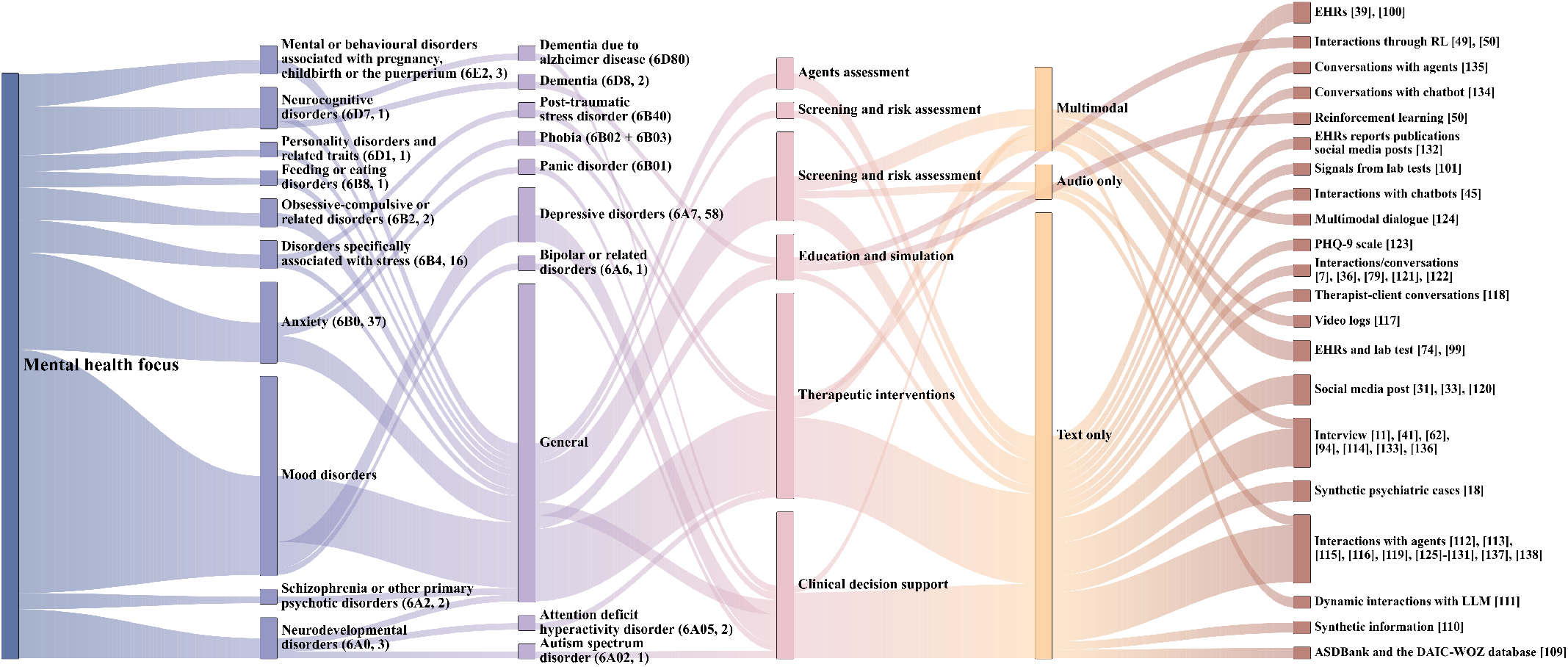
Sankey diagram linking mental health focus, downstream task type, input modality and evidence source in the surveyed literature. Left to right, flows originate from ICD-11 diagnostic groupings (for example, mood disorders, anxiety, disorders specifically associated with stress, neurodevelopmental and neurocognitive disorders) and their subtypes (for example, depressive disorders, bipolar disorders, post-traumatic stress disorder, autism spectrum disorder). These are mapped to downstream tasks (clinical decision support, therapeutic interventions, screening and risk assessment, education and simulation, agent assessment), then to dominant modalities (text only, audio only, multimodal) and finally to representative data sources (for example, interviews, therapist–client conversations, social media posts, electronic health records and lab tests, synthetic psychiatric cases and interactions with agents or chatbots). Individual studies may contribute to more than one flow when they target multiple disorders, tasks or data sources.

Within this disorder-focused subset, work on depression and anxiety is comparatively mature and spans different levels of clinical specificity and evidence. At a more general level, one study analyzes dialogues from user interactions with an assistant chatbot to classify and detect anxiety and depression, offering users a broad, holistic perspective on their mental state prior to formal health care engagement [119]. Other studies pursue more detailed and physiology-informed approaches. For example, functional near-infrared spectroscopy (fNIRS), an emerging optical neuroimaging technology, has been used to extract features from diverse brain regions that are then leveraged to assist in diagnosing anxiety and anxiety-related depression and to support more intelligent treatment planning [78]. To improve intervention quality, one chatbot design emphasizes high social cues by incorporating emotional tone modulation, facial animations and body language, combined with the integration of the GAD-7 and PHQ-9 scales. This high–social-cue system achieves better effectiveness in alleviating depression and anxiety symptoms than a low–social-cue counterpart [143]. Together, these examples illustrate how depressive and anxiety disorders have become a testbed for exploring richer sensing, multimodal inputs and experimentally controlled evaluations.

Beyond depression and anxiety, a smaller but growing body of work targets more acute or specialized conditions. For suicidality, recent studies use theoretical frameworks such as the interpersonal theory of suicide to evaluate LLM-based crisis agents and to analyze how well their responses align with clinical understandings of suicide risk and protective factors [74]. In neurodevelopmental disorders, two recent works focus on attention-deficit/hyperactivity disorder (ADHD): one examines how students might misuse ChatGPT to feign ADHD symptoms in self-report measures [114], and another reports qualitative insights from therapists and patients on integrating AI into ADHD therapy workflows [115]. These studies highlight both opportunities, for example, support in psychoeducation and coaching and novel risks, for example, gaming assessments when AI agents are introduced into neurodevelopmental care.

For severe and chronic conditions, coverage remains thin. A few works explore dementia and cognitive impairment, for instance using chess-based cognitive training programs supported by AI agents to maintain or improve mental abilities in older adults [53]. Substance-related and addictive disorders are even more sparsely represented, with early efforts focusing on AI-based chatbots to support addiction treatment and on the psychological and ethical factors that may shape uptake of AI-augmented care in these populations [144]. Sleep–wake disorders [145], [146], eating disorders and complex comorbid presentations appear only incidentally, often as inclusion criteria in broader “mental health” studies rather than as primary targets in system design.

Overall, the current distribution of mental health foci reflects both pragmatic and scientific considerations: depression and anxiety are highly prevalent (22.25% and 16.99%, respectively), more socially acceptable to discuss in digital settings, and more frequently represented in available datasets, which makes them natural early targets for AI agents. However, from a clinical perspective, the relative neglect of severe mental illness, neurocognitive disorders (0.24%), substance use (0.96%) and other high-risk conditions is a critical blind spot. Agents designed and evaluated primarily on mild-to-moderate depression and anxiety may not adequately address (and could unintentionally exacerbate) risks in populations with psychosis, bipolar disorder, suicidality, complex trauma or multi-morbidity. In later sections, we return to this mismatch between topical coverage and real-world burden of disease, and we argue that future work should explicitly prioritize under-served diagnostic groups, co-occurring conditions and high-acuity settings when developing and evaluating mental health agents.

## Demographics: age, geography, and sex as prerequisites for equitable personalization

Demographic reporting in the reviewed systems is normalized along three axes: age group, geography, and sex. Across these dimensions, we observe a recurring pattern: many studies either omit key demographic descriptors or provide them only in aggregate form, even when the intended use case implies personalization. This gap has direct implications for equity and safety. Without clearly documented age, regional, and sex distributions, it becomes difficult to judge whether an apparently strong result reflects broad clinical generalizability or is instead tightly coupled to a narrow, demographically homogeneous sample.

As summarized in Figure 5, most studies in our corpus do not specify age group at all, often because they rely on fully synthetic datasets or because privacy protections are im-plemented by removing or coarsening demographic attributes. Among the studies that do report age, a substantial proportion focus on adolescents and young adults, especially in school and university contexts. This emphasis cuts across different methodological paradigms. For example, Ni et al. simulate school counseling sessions to evaluate both counseling skill and the risk of unsafe or inappropriate information delivery in a controlled, youth-oriented setting [147]. Other work targets scalable support for postsecondary students: Luna is designed as a campus-facing agent that combines LLM-based conversation with safety guardrails and referral logic to support ethical, large-scale deployment in university environments [148]. In youth service settings, the Frontline Assistant: Issue Identification and Recommendation (FAIIR) uses domain-adapted transformer models to improve issue detection for help-seeking young people, explicitly positioning itself as an upstream triage tool rather than a replacement for therapy [149]. PsyConnect similarly illustrates a student-facing design, using an AI-powered platform to infer emotional states from text and provide monitoring and support for stress, anxiety, and depression in academic environments [150]. Together, these systems demonstrate how age-targeted designs can be tailored to educational settings, but they also highlight a relative lack of work on older adults, pediatric populations outside school settings, and life stages such as midlife or late-life where the mental health burden is also substantial.

**Fig. 5.**
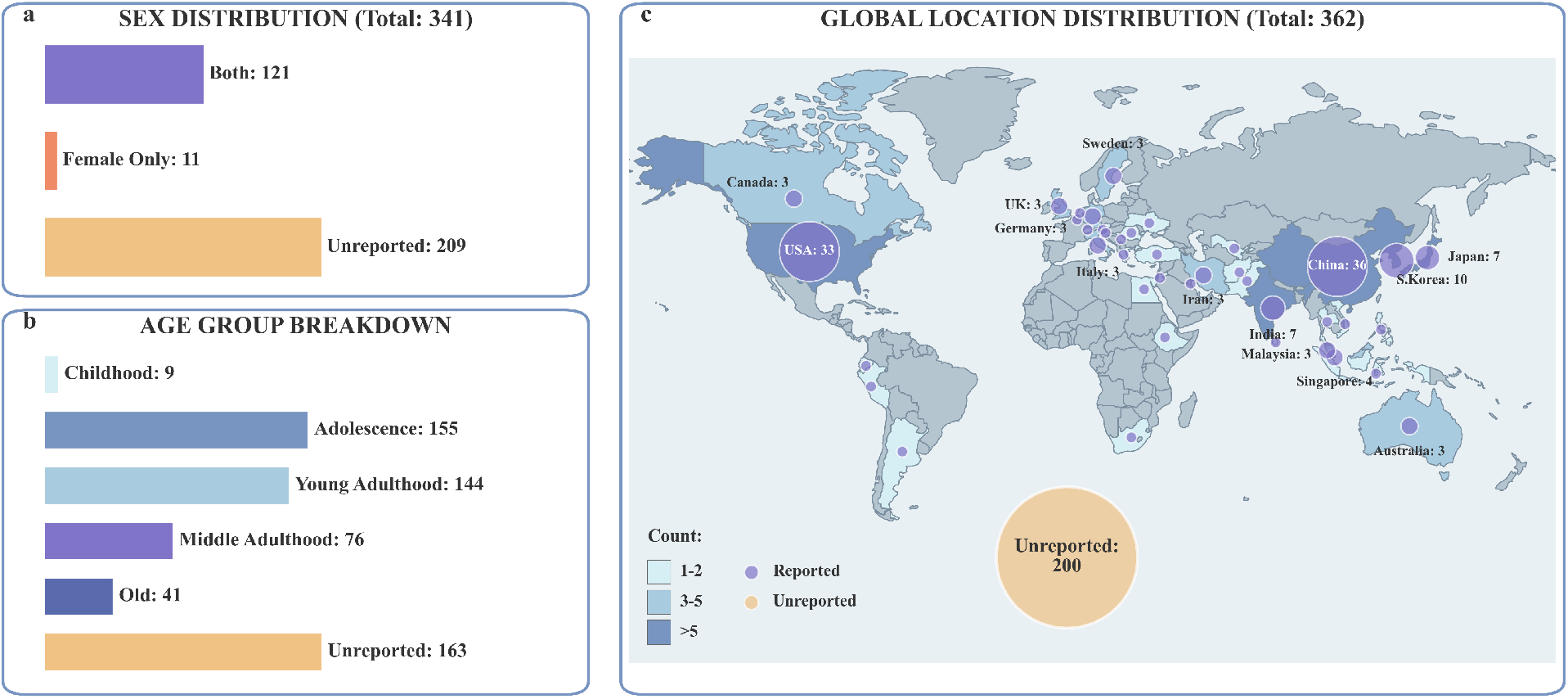
Demographic reporting across the current landscape of AI in mental health agents. (a) Sex distribution for studies that reported participant sex (both male and female, female only) versus those that did not report this information. (b) Age-group breakdown, showing counts for childhood, adolescence, young adulthood, middle adulthood and older adulthood, alongside the number of studies with unspecified age. (c) Global study-location distribution, with bubble size indicating the number of studies conducted in each country and a large aggregate bubble for studies that did not report location.

With respect to sex, most studies that report this variable include both male and female participants, but disorder-specific systems often concentrate on populations where sex-linked risk is particularly salient. This is especially apparent in work on postpartum mental health. Several studies design conversational agents specifically for postpartum people, using context-sensitive rule-based or generative chatbots to deliver human-like, empathetic responses tailored to postpartum depression and anxiety [141], [142]. Beyond conversational support, ClinPreAI exemplifies how agentic systems can incorporate multimodal electronic health record data to predict postpartum depression risk, illustrating that postpartum-focused research now spans both interactive support and clinically oriented risk modeling [36]. At the same time, there is relatively little work that explicitly reports or optimizes for nonbinary or gender-diverse users, despite growing evidence that gender minority groups face disproportionate mental health burdens. Overall, sex is more often treated as a covariate in prediction models than as a basis for systematically stratified evaluation or tailored intervention logic.

Geographic and cultural coverage show similar imbalances. Among studies that explicitly report location, 33 were conducted in the USA and 36 in China, indicating a concentration of evidence in North America and East Asia. Far fewer studies originate from low- and middle-income countries (LMICs) or from regions where digital mental health infrastructures are still emerging. Yet there are important exceptions that point toward more globally inclusive trajectories. In Sri Lanka, a mixed-methods study on engagement of medical practitioners with an AI-powered mental health decision-support chatbot examines technological, organizational, and environmental factors that shape adoption, and evaluates a prototype system built on the Microsoft Azure platform [151]. This work illustrates how local resource constraints and governance structures can shape both attitudes toward AI and the practical feasibility of deployment.

Cross-cultural preference and trust studies also begin to delineate how attitudes toward AI-mediated care vary across settings. In a large online vignette experiment with 1,183 participants from German-speaking countries (Germany, Austria, Switzerland), Riedl et al. compare hypothetical consultations with a human doctor, a doctor supported by AI, and an AI chatbot alone. Participants consistently preferred the human doctor, followed by doctor-plus-AI, with the AI-only condition rated lowest on trust, willingness to disclose information, treatment adherence, and satisfaction [152]. This pattern underscores that even in high-resource settings with strong digital infrastructures, AI chatbots are not yet perceived as acceptable stand-alone replacements for clinicians, especially in sensitive domains such as psychiatry. Parallel lines of work on cultural adaptation further suggest that baseline LLM outputs often reflect Western conversational norms, and that explicit cultural prompting or localization is needed to improve perceived empathy and relevance for users from non-Western backgrounds [153]. Beyond the Anglophone and East Asian contexts, PsychoLexTherapy proposes a structured framework for simulating psychotherapeutic reasoning in Persian, emphasizing privacy-preserving, culturally aligned reasoning steps rather than direct end-user deployment, and thereby opening a path toward culturally grounded agent design in under-served languages [154].

Taken together, these observations indicate that demographic representation is not merely a reporting detail but a precondition for equitable personalization. Age, geography, and sex distributions shape both the linguistic content and the clinical phenotypes that models encounter during training and evaluation. For future work, we identify three concrete directions. First, systems should routinely report demographic breakdowns and, where feasible, stratified performance metrics, particularly when claims of personalization or fairness are made. Second, there is a need for prospective studies that explicitly recruit underrepresented groups, including older adults, gender-diverse individuals, and communities in LMICs, and evaluate whether agentic systems maintain safety and effectiveness across these strata. Third, cross-cultural adaptation should be treated as a core design concern rather than an afterthought, with locally grounded co-design processes and language-specific reasoning frameworks (as in Persian or other non-English settings) used to align AI behavior with local values, communication styles, and care pathways across diverse regions.

## Downstream tasks: mapping claims to interfaces, workflows, and evidence needs

In our framework, the *downstream task* dimension describes what mental health agents are actually used for and how these uses are embedded into clinical or everyday workflows. We organize tasks into six broad categories: (i) screening and triage, (i) clinical decision support, (iii) therapeutic interventions, (iv) documentation generation, (v) ethical and legal support and (vi) education and simulation. Each task category is associated with characteristic interfaces, for example, chat-based versus non-chat, workflow insertion points, for example, patient-facing versus clinician-facing versus background services and distinct evidence requirements, for example, diagnostic accuracy versus symptom reduction versus usability and trust. To clarify the distinct computational demands of the first three core tasks, we explicitly formalize their learning objectives below.

### Screening and triage

Screening and triage tasks focus on identifying potential mental health disorders early and routing users to appropriate levels of care. Computationally, this corresponds to a **Static Classification** problem where the system learns a mapping function *h*_*ω*_ from input space 𝒳 to *N*_*c*_ risk classes {0, …, *N*_*c*_} to minimize a prediction loss ℒ_*pred*_ against ground-truth labels *y*:

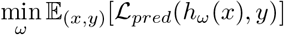

Screening systems typically estimate symptom severity or risk and can trigger different follow-up actions, such as self-help recommendations, referral to a clinician or emergency escalation. In AI-based settings, screening and triage may be implemented via short conversational assessments [76], video and audio analysis [121] or standardized scales integrated directly into dialogue [48]. For example, one line of work describes passive tools that continuously screen text-based media or message streams for emotional tone and risk markers, enabling background crisis detection and triage without an explicit conversational user interface [65]. Other studies use audio features alone or in combination with language to estimate depression risk, providing risk scores that can be monitored over time [66]. MAQuA [29] proposes an adaptive question-asking framework for simultaneous, multi-dimensional mental health screening: by dynamically selecting the most informative next questions across multiple symptom dimensions, MAQuA aims to streamline data collection and accelerate both screening and diagnostic workflows. Despite this variety, relatively few systems report classical diagnostic metrics, for example, sensitivity, F1 Score, specificity, Positive Predictive Value and Negative Predictive Value [44], against clinician-verified labels; many instead rely on proxy outcomes such as self-report agreement or LLM-based judgments, which weakens the evidentiary basis for deployment as clinical triage tools.

### Evidence synthesis: diagnostic accuracy meta-analysis

To quantify the diagnostic performance of AI-based screening and triage systems, we conducted a systematic meta-analysis covering 11 study records from 9 independent studies, comprising a combined *N* = 147,780 assessments. Studies spanned a broad spectrum of conditions: generalised anxiety [21], mood disorders including depression and bipolar disorder [22], neurodevelopmental conditions including aphasia, ASD and depression classification [113], postpartum depression [36], youth mental-health crisis identification [149], OCD [137], suicide risk stratification [82], audio-based depression screening [66], and anxiety–depression classification [119]. A DerSimonian–Laird random-effects model was applied separately to logit-transformed sensitivity and specificity; methodological quality was assessed with the QUADAS-2 framework (Table IV).

**TABLE IV.**
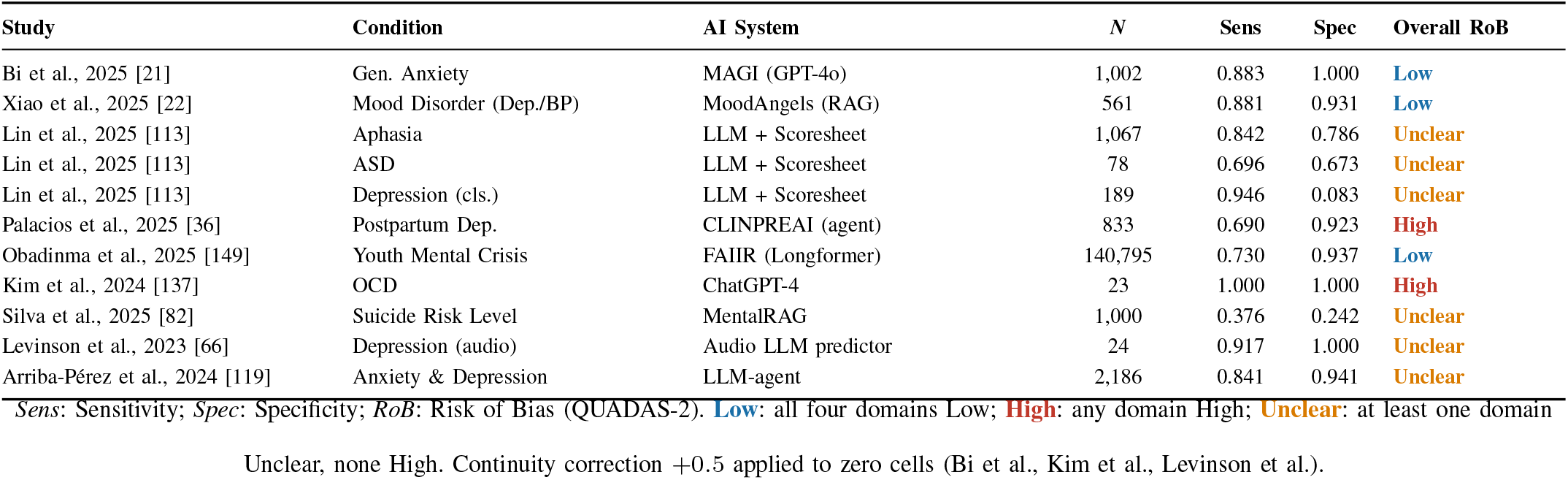
Characteristics and QUADAS-2 Overall Risk-of-Bias of Studies Included in the Diagnostic-Accuracy Meta-Analysis (*n* = 11 records from 9 studies; *n* = 147,780 assessments)

Pooled across all 11 records, AI-based diagnostic agents achieved a sensitivity of 0.799 (95% CI: 0.711–0.866) and a specificity of 0.848 (95% CI: 0.668–0.939), with a Diagnostic Odds Ratio (DOR) of 39.7 (Table V; Figures 6–8 Substantial heterogeneity was present in both estimates (*I*^2^ = 97.4% for sensitivity; *I*^2^ = 99.5% for specificity), attributable to the diversity of psychiatric conditions, variability in reference standards (from licensed clinician consensus to self-report scale cut-offs) and architectural heterogeneity across fine-tuned transformers and multi-agent GPT-4 interview systems. A pre-specified sensitivity analysis restricted to the three QUADAS-2 Low-Risk studies (combined *N* = 142,358) substantially raised specificity to 0.944 (95% CI: 0.908–0.967) and increased the DOR nine-fold to 362.2 (Figure 9), confirming that methodological rigour is a key moderator of apparent performance and supporting clinical utility as a *rule-in* screening tool in human-in-the-loop workflows. Two records illustrate performance boundaries: Kim et al. [137] achieved perfect classification (sensitivity and specificity both 1.00) on *N* = 23 published OCD clinical vignettes—a known optimistic-bias artifact from spectrum-selected cases—while the fine-grained suicide risk-level prediction task of Mental-RAG [82] yielded the lowest overall performance (sensitivity 0.376, specificity 0.242), underscoring the importance of consecutive enrollment and clinician-verified ground-truth labels for valid accuracy benchmarking.

**TABLE V.** Pooled Diagnostic Accuracy: Primary Analysis vs. Low-Risk Sensitivity Analysis.

|  |  |  |  |  |
| --- | --- | --- | --- | --- |
| Analysis | *n* | Sensitivity (95% CI) | Specificity (95% CI) | DOR |
| All studies | 11 | 0.799 (0.711–0.866) | 0.848 (0.668–0.939) | 39.7 |
| *I*<sup>2</sup> |  | 97.4% | 99.5% | — |
| Low-Risk only | 3 | 0.834 (0.687–0.920) | 0.944 (0.908–0.967) | 362.2 |
| *I*<sup>2</sup> |  | 92.1% | 82.5% | — |

DerSimonian–Laird random-effects on logit-transformed estimates. DOR: Diagnostic Odds Ratio. Low-Risk subset (*N* = 142,358): Bi et al. 2025, Xiao et al. 2025, Obadinma et al. 2025.

**Fig. 6.**
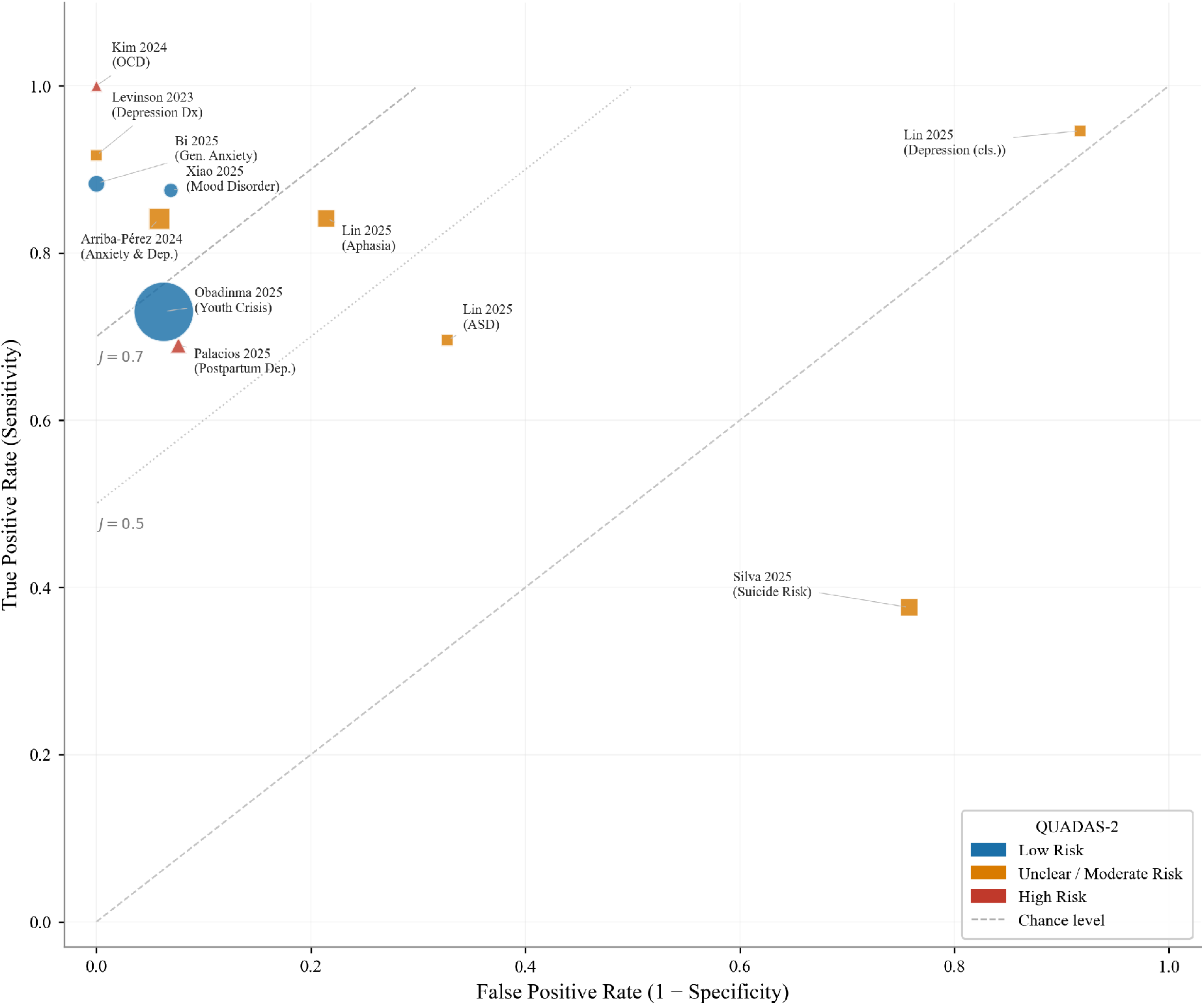
Summary ROC (SROC) space for diagnostic-accuracy meta-analysis (*n* = 11 records; *N* = 147,780). Each marker represents one study record; marker area is proportional to 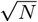; shape and color encode the QUADAS-2 overall risk of bias (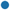 Low, 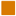 Unclear, 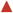 High). Iso-accuracy lines show Youden’s Index *J* = 0.7 (dashed) and *J* = 0.5 (dotted). Large-sample studies cluster in the upper-left region of high sensitivity and high specificity.

**Fig. 7.**
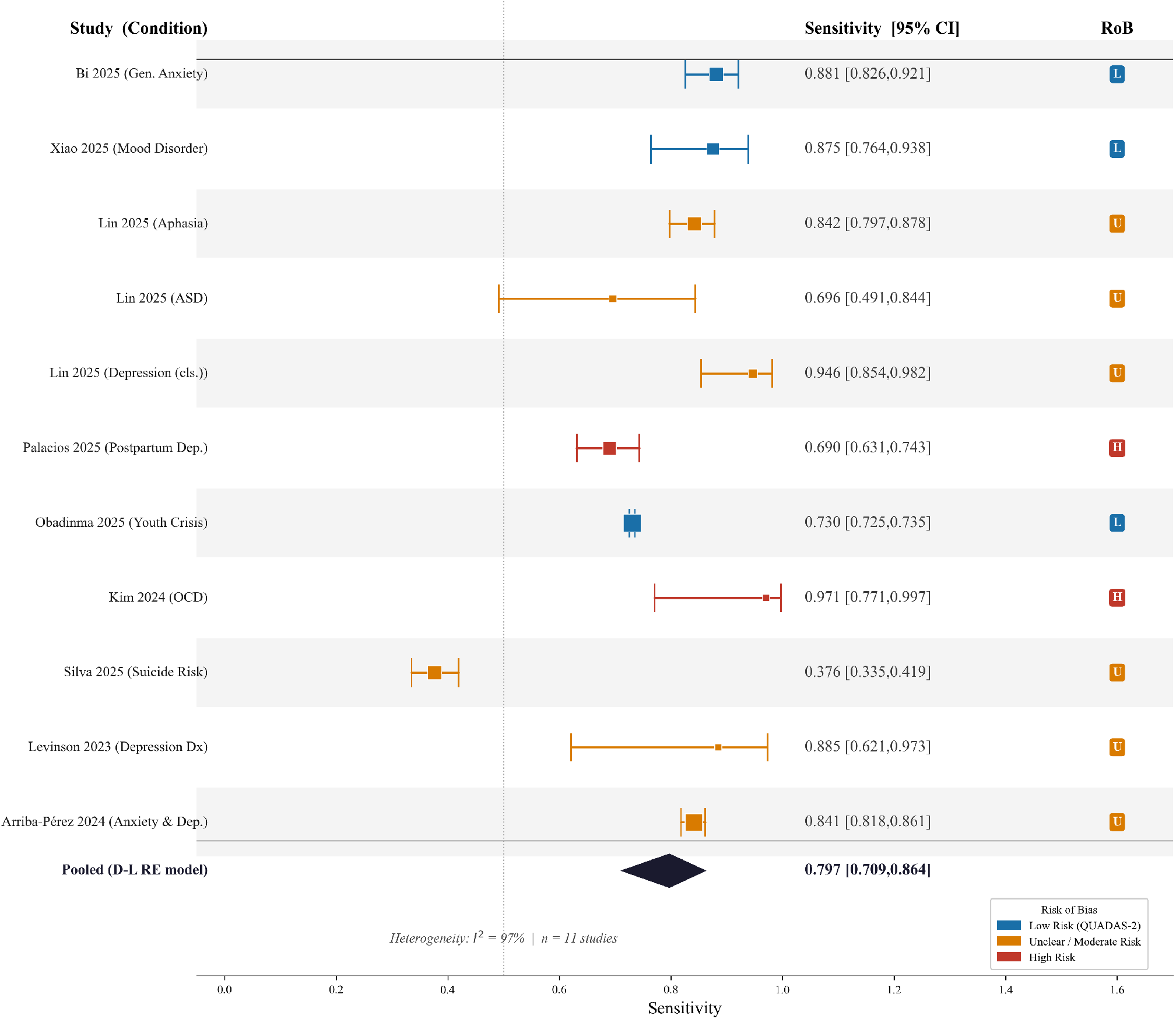
Forest plot of sensitivity. Squares indicate per-study point estimates with area proportional to 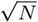; horizontal bars are 95% Wilson confidence intervals. Color encodes QUADAS-2 risk of bias. The diamond shows the DerSimonian–Laird pooled estimate: 0.799 (95% CI: 0.711–0.866); *I*^2^ = 97.4%.

**Fig. 8.**
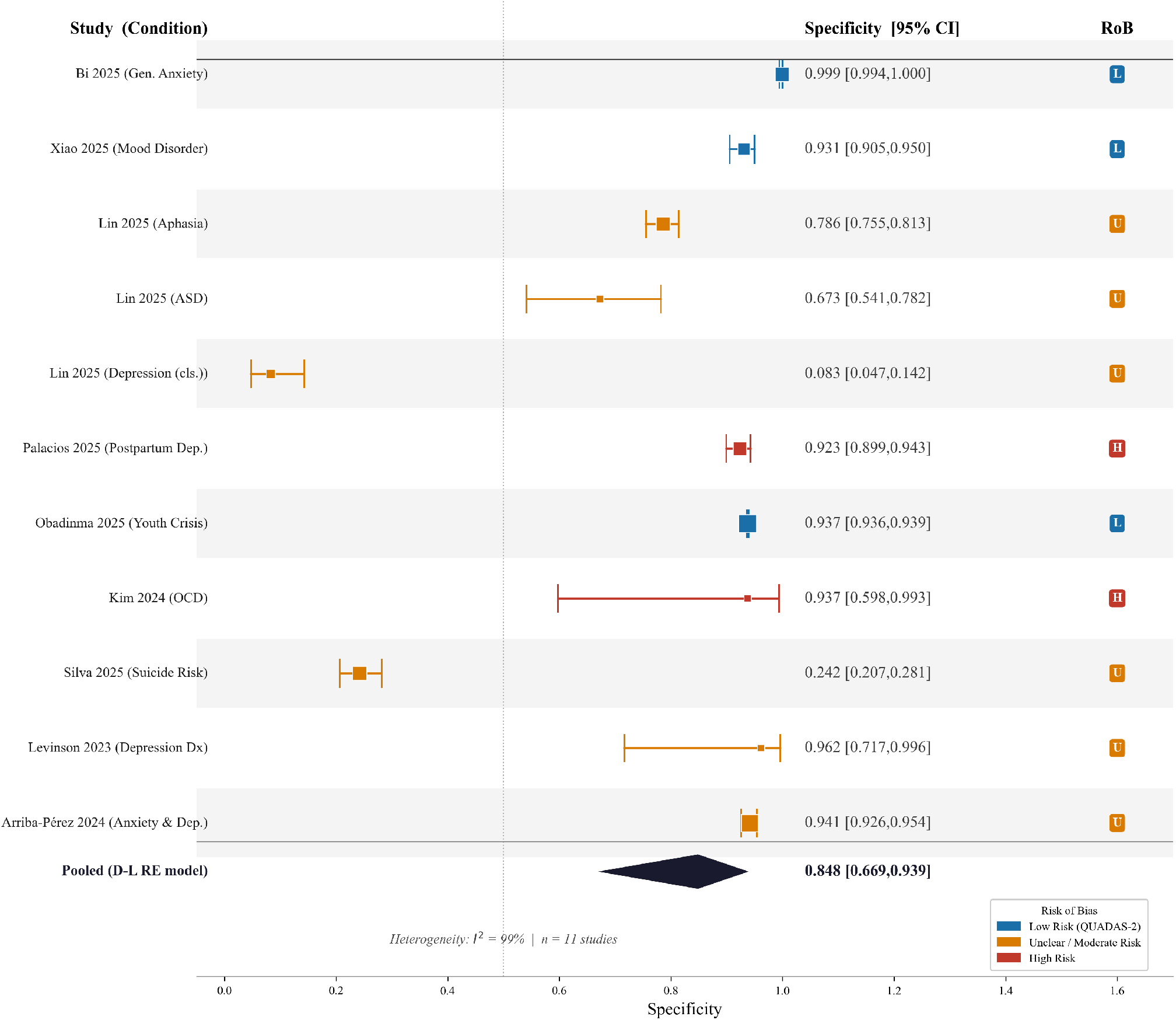
Forest plot — specificity (screening-and-triage diagnostic-accuracy synthesis). Squares indicate per-study point estimates with area proportional to 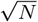; horizontal bars are 95% Wilson confidence intervals; color encodes QUADAS-2 risk of bias. Pooled specificity: 0.848 (95% CI: 0.668–0.939); *I*^2^ = 99.5%. The marked improvement among Low-Risk studies (0.944 vs. 0.848) signals a systematic influence of methodological quality on specificity estimates.

**Fig. 9.**
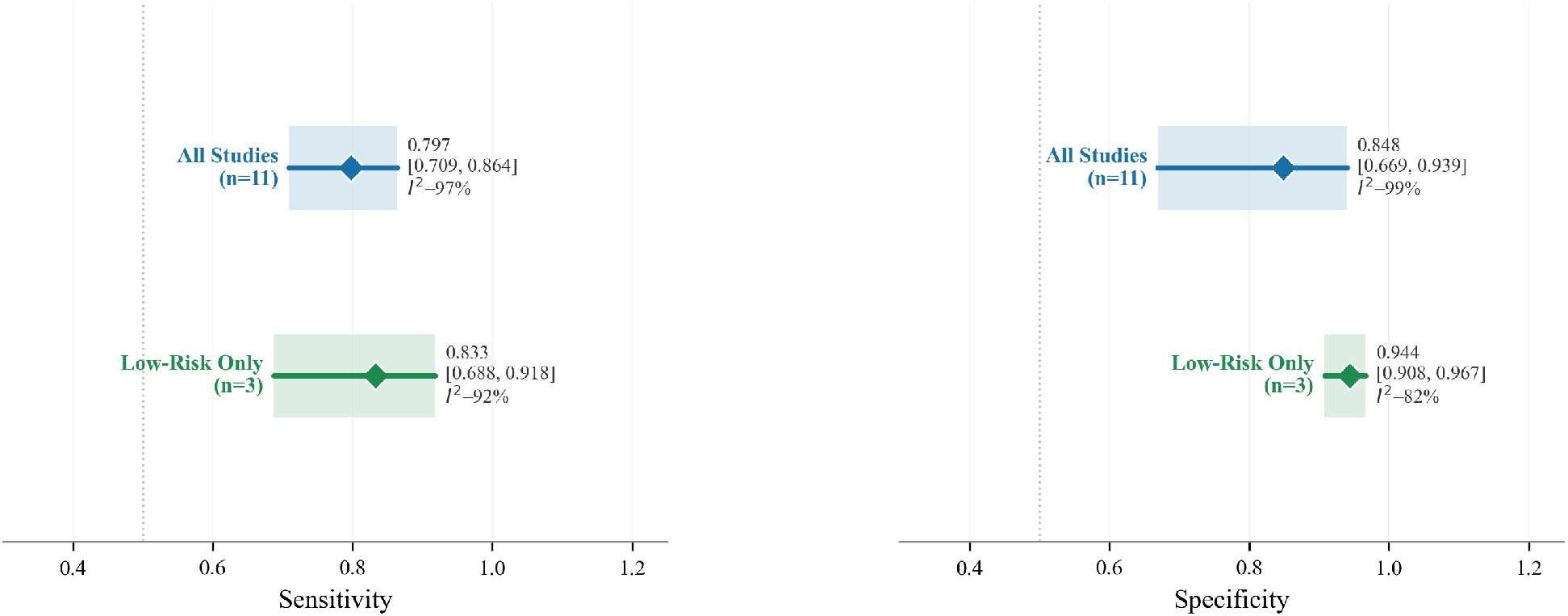
Sensitivity analysis: all studies vs. Low-Risk subset. Diamonds denote pooled estimates with 95% CIs. Restricting to Low-Risk studies (QUADAS-2 all domains Low) raises specificity from 0.848 to 0.944 and increases the DOR nine-fold from 40 to 362, confirming methodological quality as a significant moderator of apparent diagnostic performance.

### Clinical decision support

Clinical decision support (CDS) tasks are designed to help clinicians make better-informed decisions, rather than to replace their expertise. Formally, this is a **Utility Maximization** task where the goal is to recommend a clinical action *a* ∈ 𝒜 (e.g., medication selection) that maximizes the expected patient utility *U* over predicted outcomes *y*_outcome_, typically derived from guidelines:

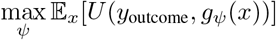

CDS systems draw on patient data, clinical guidelines and prior cases to assist with assessment, diagnosis, treatment planning, ongoing monitoring and documentation. Some agents explicitly position themselves as “AI psychiatrists” that can propose differential diagnoses or severity ratings based on conversational or structured inputs [22], [83]. Others focus on narrow decision points, such as assessing treatment response or predicting relapse risk using longitudinal data from electronic health records or patient-reported outcomes [36], [104]. Multi-agent frameworks like MoodAngels [22] allocate diagnostic roles to several agents and then aggregate their opinions through judge and debate mechanisms to improve robustness. Visual reasoning agents have also been proposed to help clinicians interpret psychometric drawings or neuroimaging outputs, supplying structured interpretations rather than raw scores [89]. In all these cases, the interface is often clinician-facing, for example, dashboards or report summaries rather than direct-to-patient chat, and appropriate evaluation requires not only predictive performance but also assessments of workflow fit, trust and potential overreliance on AI recommendations.

### Therapeutic interventions

Therapeutic intervention tasks aim to deliver or augment elements of psychotherapy and counseling, rather than merely screening or providing information. Unlike the single-step tasks above, therapy is modeled as a **Sequential Optimization** problem. Reusing the agent notation from Section II, the objective is to learn a policy *π* that maximizes the cumulative reward *r* (encoding symptom relief or alliance) over a trajectory, discounted by a factor *η*:

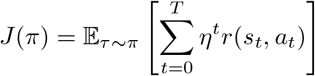

Systems in this category help users regulate emotions, challenge maladaptive thoughts, practice coping strategies or build behavioral routines using approaches such as cognitive–behavioral therapy (CBT), problem-solving therapy and motivational interviewing. AutoCBT [61] exemplifies a multiagent CBT framework in which different agents take on roles such as reformulating user statements, suggesting cognitive restructuring strategies and evaluating the quality of generated interventions; the system is evaluated on its adherence to CBT techniques and perceived usefulness. Other work uses LLMs to power CBT-style conversations and examines how different disclosure levels and prompt designs affect user engagement and perceived effectiveness [155]. Except for building the CBT itself, one study further utilized machine learning techniques to predict user adherence to digital cognitive behavioral therapy for insomnia (dCBT-I) [156], which can allow clinicians to tailor sleep-focused treatments based on the observed and predicted adherence patterns. PsychoGAT [91] provides a generic gamified framework for psychological assessment and intervention by integrating standardized scales into personalized fiction games, where users’ in-game choices and interactions with designated roles are used to infer mental health attributes and tailor feedback. Additional examples include motivational-interviewing agents that simulate therapist behaviors over multi-turn dialogues [16], [93] and social-support agents that provide empathic responses grounded in personality or inter-personal context [20]. For therapeutic tasks, the evidentiary bar is particularly high: beyond self-report satisfaction or short-term symptom changes, rigorous evaluation will ultimately require controlled trials and long-term follow-up.

### Evidence synthesis: clinical symptom improvement meta-analysis

To provide a pooled effectiveness estimate across the intervention studies in our corpus, we conducted a random-effects meta-analysis aggregating 13 outcome measures from studies reporting validated psychological symptom scales, including the PHQ-9, GAD-7, DASS-21, GHQ-28, PSS, BAI, and BFNE-II. The majority of individual effect sizes were negative, indicating reductions in symptom scores following AI-based mental-health interventions. The pooled random-effects estimate confirmed a statistically significant small-to-moderate improvement (Hedges’ *g* = −0.383, 95% CI [−0.591, −0.174]; Table VI; Figure 10), with negative values encoding symptom reduction. Substantial between-study heterogeneity was present (*Q* = 76.15, *df* = 12, *I*^2^ = 84.24%, *τ* ^2^ = 0.112), likely reflecting variability in intervention design, underlying model architecture, treatment duration, outcome instruments and population characteristics. These findings confirm that AI-driven therapeutic agents can produce clinically detectable symptom improvements. However, the high heterogeneity underscores that effect magnitude varies considerably across deployed systems: the pooled estimate should be interpreted as a summary tendency rather than a universal benchmark, reinforcing the need for head-to-head comparative trials and standardised outcome reporting in future work.

**TABLE VI.**
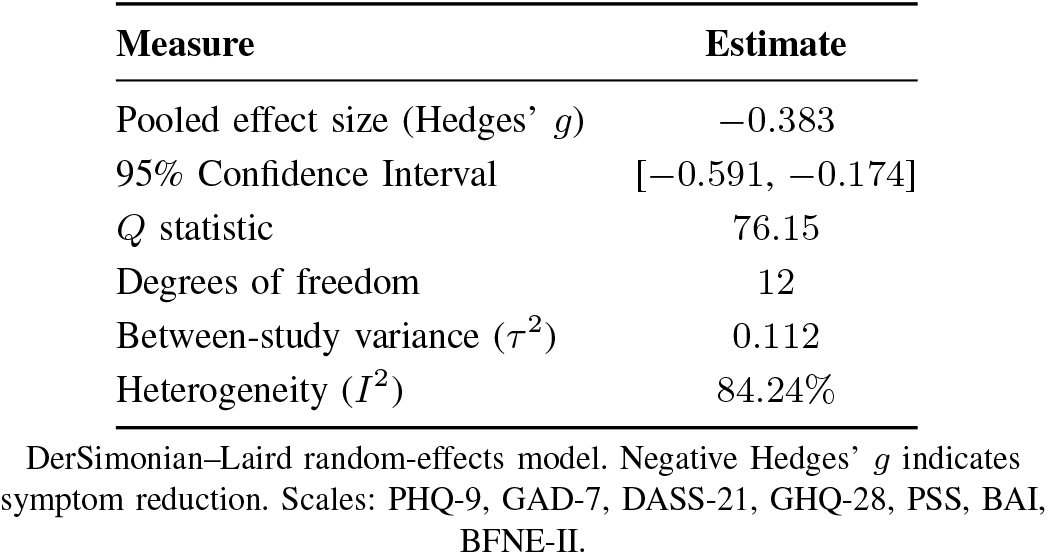
Random-Effects Meta-Analysis Results for Clinical Symptom Improvement (*k* = 13 outcome measures)

|  |  |
| --- | --- |
| Measure | Estimate |
| Pooled effect size (Hedges' *g*) | −0.383 |
| 95% Confidence Interval | [−0.591, −0.174] |
| *Q* statistic | 76.15 |
| Degrees of freedom | 12 |
| Between-study variance ($\tau^2$) | 0.112 |
| Heterogeneity (*I*<sup>2</sup>) | 84.24% |

DerSimonian–Laird random-effects model. Negative Hedges' *g* indicates symptom reduction. Scales: PHQ-9, GAD-7, DASS-21, GHQ-28, PSS, BAI, BFNE-II.

**Fig. 10.**
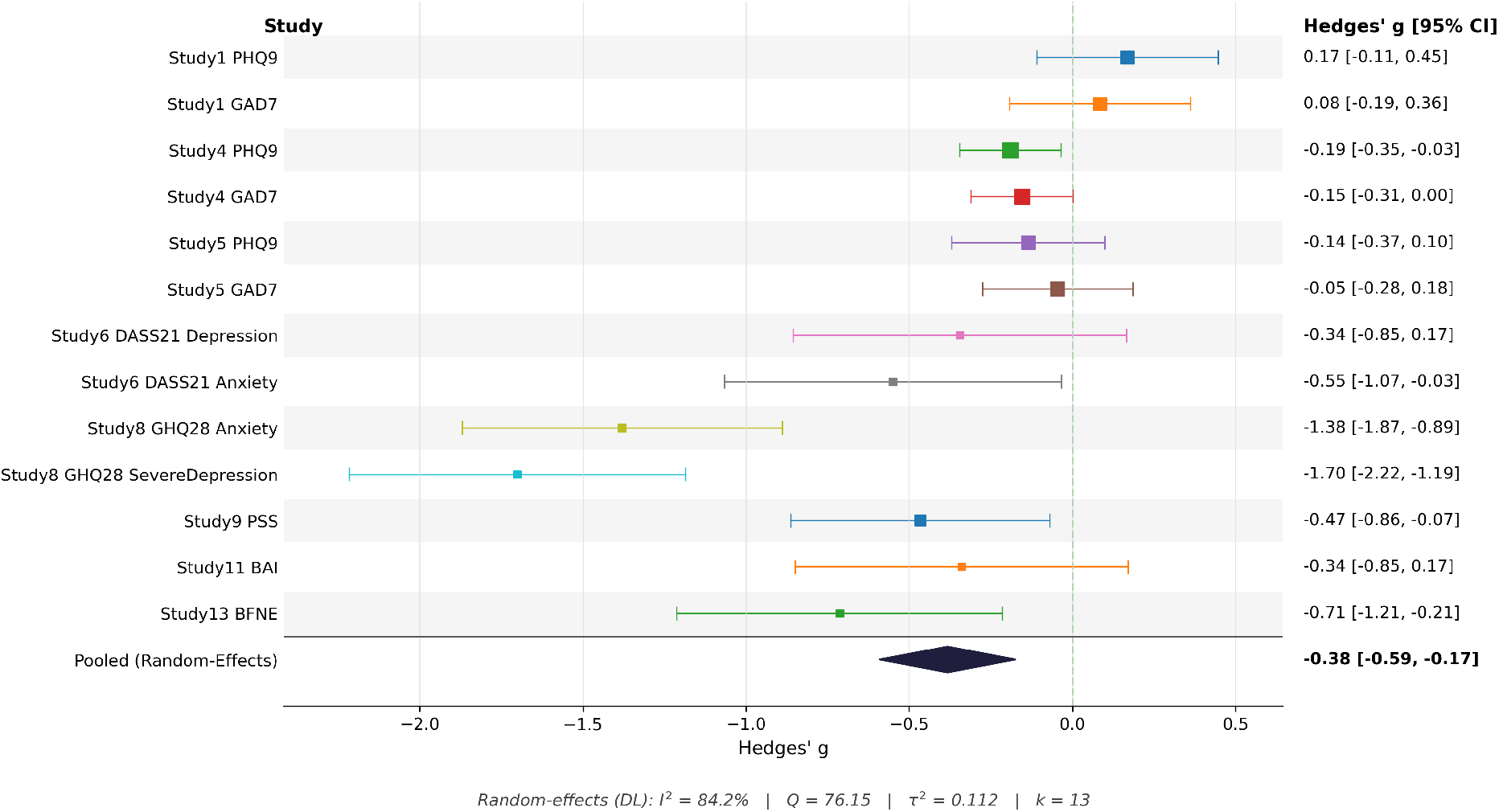
Forest plot for meta-analysis of clinical symptom improvement (*k* = 13 outcome measures across validated psychological scales). Negative effect sizes indicate symptom reduction following AI-based mental-health interventions. The pooled random-effects estimate (diamond) is Hedges’ *g* = *−*0.383 (95% CI [*−*0.591, *−*0.174]); *I*^2^ = 84.24% reflects substantial heterogeneity across intervention designs, populations and outcome instruments.

### Documentation generation

Documentation generation tasks focus on helping clinicians produce structured clinical documents such as progress notes, intake summaries or test result reports. Computationally, this is modeled as a Con-strained Summarization task. The system maps an unstructured interaction history ℋ to a structured report *d*. Unlike openended chat, the objective function includes a strict penalty for extrinsic hallucinations (information not present in source ℋ), weighted by a coefficient *λ*:

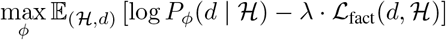

Rather than typing everything from scratch, AI agent systems assist clinicians to draft notes and then edit them, potentially reducing documentation burden and freeing time for direct patient care. Multi-agent dialogue systems can, for example, transform raw conversations into structured intake summaries by extracting key symptoms, histories and risk factors [69]. PsyDraw [89] adopts four agents to capture and analyze drawing features in parallel at the overall, house, tree and person levels, then passes the extracted features to a second-stage set of agents that generate narrative reports for therapists to interpret. AliceBot [132] generates longitudinal mental health reports based on sentiment and topic trajectories inferred from conversational interactions, providing users and clinicians with a summary of emotional patterns over time. Other decision-support systems generate draft diagnostic impressions or treatment plans from EHR data and conversational context [83], [104]. For documentation tasks, key outcomes include factual accuracy, preservation of clinically critical details, reduction in documentation time and the extent to which clinicians remain appropriately “in the loop” as final arbiters of record content.

### Ethical, legal and safety support

Ethical and legal support tasks aim to ensure that mental health AI agent systems behave safely, respect user rights and comply with regulations. In our corpus, most such efforts focus on data governance: systems protect privacy through anonymization [80], deliberate de-identification using named entity recognition and reversible pseudonymization [71], [157] or careful curation of training datasets to exclude sensitive content [121]. A smaller but growing body of work targets the *behavior* of models themselves. Safety evaluation frameworks such as PsySafe [76] use multi-agent setups to probe how interacting agents may amplify or dampen risky behaviors in high-stakes scenarios, including self-harm and suicide risk. Other systems introduce dedicated “safety critic” or “refiner” agents that sit between the core model and the user, filtering, editing or rejecting potentially harmful responses [8], [75]. Design-oriented studies also examine factors such as transparency, explainability and recourse (for example, how users can contest or correct model outputs) in counseling or triage settings [151], [158]. Overall, however, explicit ethical and legal support features are still more often described at a high level than evaluated empirically, and few systems undergo formal audits or alignment assessments grounded in mental health specific regulatory guidance.

### Education and simulation

Finally, a distinct cluster of systems uses AI agents to support education and simulation for clinicians, trainees or peer supporters. These systems typically rely on multi-agent role-play or virtual standardized patients to create realistic, controllable training scenarios. MAGI [21], for example, is designed for psychiatric assessment interviews: multiple agents take on roles such as interviewer, judgment and diagnosis, enabling trainees to practice the flow of structured assessments and receive feedback. CAMI [16] uses a client simulator and counselor agent to model motivational interviewing sessions, providing a platform for both practicing and evaluating counseling skills. ESC-Judge [23] simulates emotional-support conversations and uses multi-stage, multiagent evaluation to assess the quality of support, offering a blueprint for training and benchmarking support agents. Other frameworks generate synthetic supervision signals by simulating expert feedback on trainee–patient interactions, which can then be used both for human education and for improving AI agents themselves [87], [88]. Educational and simulation systems are typically evaluated on the realism of generated interactions, alignment with established therapeutic techniques and their impact on trainee performance, rather than direct patient outcomes, sharing a similar set of challenges and opportunities with the general LLM-based agents [159]. Nonetheless, because they shape how future clinicians reason about and interact with patients, their design and evaluation have important downstream implications for clinical practice.

Across all these task categories, a key theme is the need to align *claims* with *evidence*. Screening and triage systems should be evaluated on clinically grounded diagnostic metrics; CDS tools should be assessed on decision quality, workflow fit and avoidance of overreliance; therapeutic agents require rigorous trials and safety monitoring; documentation assistants must demonstrate factual accuracy and time savings without eroding clinician oversight; ethical and legal modules need systematic auditing; education and simulation platforms should show measurable benefits for learner competence and patient safety. In later sections, we return to these evidence gaps and propose an evaluation ladder tailored to mental health agents.

## Evaluation: from offline metrics to clinician-and patient-in-the-loop validation

The *evaluation* dimension describes how mental health agents are assessed, and how closely those assessments approximate their intended real-world use. Across our corpus, evaluations can be broadly grouped into (i) automated or offline metrics, (ii) human evaluations by clinicians or lay users and (iii) emerging benchmark frameworks that combine synthetic users, safety stress tests and structured clinical rubrics. Taken together, these approaches allow us to assess system performance and examine whether outputs align with human preferences and clinical requirements. In addition, benchmark-based evaluation provides a shared reference point for continuously tracking progress as new systems are proposed and evaluated over time.

### Automated evaluation: task performance and language quality

Within automated evaluation, studies typically follow two main routes. The first route assesses task applicability using quantitative performance indicators such as accuracy, F1 score and area under the receiver operating characteristic curve (AUROC) on classification or prediction tasks [157], [160], [161]. For example, MemoryBank-style agents and counselor-support systems report predictive metrics for tasks such as risk classification, emotion recognition and outcome prediction, often on held-out datasets or cross-validation splits. ChatCounselor-type systems additionally report sensitivity and specificity for detecting high-risk cases in simulated triage workflows [157].

The second route evaluates language quality using standard text-generation benchmarks such as BLEU and ROUGE, particularly for empathetic or supportive dialogue generation [86], [162]. In these settings, model outputs are compared against reference responses from human counselors or crowdworkers. Some studies also use diversity metrics, for example, distinct-*n*, and perplexity to capture linguistic richness and fluency [162]. Although these metrics are convenient and reproducible, they only partially approximate clinically relevant qualities such as appropriateness, empathy, or adherence to therapeutic protocols, and they are often computed on short, decontextualized snippets rather than on full multi-turn interactions.

#### Human evaluation: clinicians, lay users and voice-based settings

Human evaluation complements automated metrics by directly assessing how system outputs are perceived and judged by people. In the strongest cases, to measure whether the system’s performance will align with real clinical use or human preferences. A subset of studies relies on clinical experts or other health professionals to evaluate system behavior, focusing on clinical appropriateness, diagnostic soundness and safety [86], [99]. For instance, some works ask psychiatrists or psychologists to rate chatbot responses on scales such as “clinically appropriate,” “potentially harmful,” or “requires escalation,” and to compare AI-generated output with expert-written responses [99]. Others use therapists to annotate dialogue segments for adherence to specific therapeutic techniques, for example, cognitive restructuring, reflective listening, and then treat these annotations as labels for further automated analysis [86]. These clinician-grounded protocols move beyond preference and provide more outcome-relevant evidence about whether systems behave in ways consistent with clinical standards.

While automated metrics focus on statistical overlap, human evaluation remains the gold standard for clinical validity. To standardize subjective assessment, we formalize the human rating process as a weighted composite score *Q*_human_ over *M* clinical criteria (for example, empathy, correctness, safety):

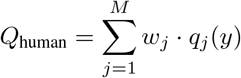

Here, *q*_*j*_(*y*) is the expert rating on the *j*-th quality dimension and *w*_*j*_ denotes its clinical-importance weight. This framing makes explicit where rater alignment, rubric design and inter-rater reliability affect overall scores.

A larger group of studies instead recruits lay participants to rate responses, typically measuring perceived helpfulness, naturalness, empathy and satisfaction [163], [164]. Such user-focused evaluations are particularly common for self-help or wellness chatbots, where immediate clinical risk is lower but long-term engagement and trust are crucial. Recent work has extended this paradigm to voice-enabled generative AI systems for mental health care, combining quantitative task metrics with user ratings of usability, rapport and perceived therapeutic alliance in spoken interactions [165]. Other safety-oriented studies evaluate general-purpose LLMs and clinical-grade chatbots in simulated clinical workflows to examine how clinicians and patients would respond to AI-generated advice [166]. Overall, human evaluations provide indispensable insight into acceptability and subjective experience, but they are often conducted on small samples and rarely involve longitudinal follow-up.

### Evidence synthesis: system usability and acceptability meta-analysis

To synthesize the fragmented evidence on user acceptance, we conducted a fixed-effect meta-analysis normalizing heterogeneous usability instruments—including the System Usability Scale (SUS), a Likert-based General Agent Rating (GAR) scale, and the Technology–Wellbeing Engagement and Emotion Scale (TWEETS)—to a unified Percentage of Maximum Possible (POMP) metric:

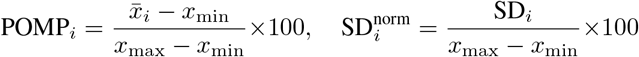

where *x*_min_ and *x*_max_ denote the theoretical minimum and maximum of each instrument (SUS: 0–100; Likert scales: 1– *k*; TWEETS: 0–30). Pooled across 12 study arms (*n*_total_ = 609), the mean POMP score was **76.9** (95% CI: 75.6–78.1; *I*^2^ = 92%, Cochran’s *Q* = 137.89, *p <* 0.001), substantially surpassing the universally recognised SUS acceptability threshold of 68 (Figure 11). This finding demonstrates broad user acceptance of LLM-driven conversational agents in psychological support contexts, despite the diversity of systems and populations evaluated. The high heterogeneity (*I*^2^ = 92%) is largely attributable to architectural and front-end design choices rather than baseline LLM capability.

**Fig. 11.**
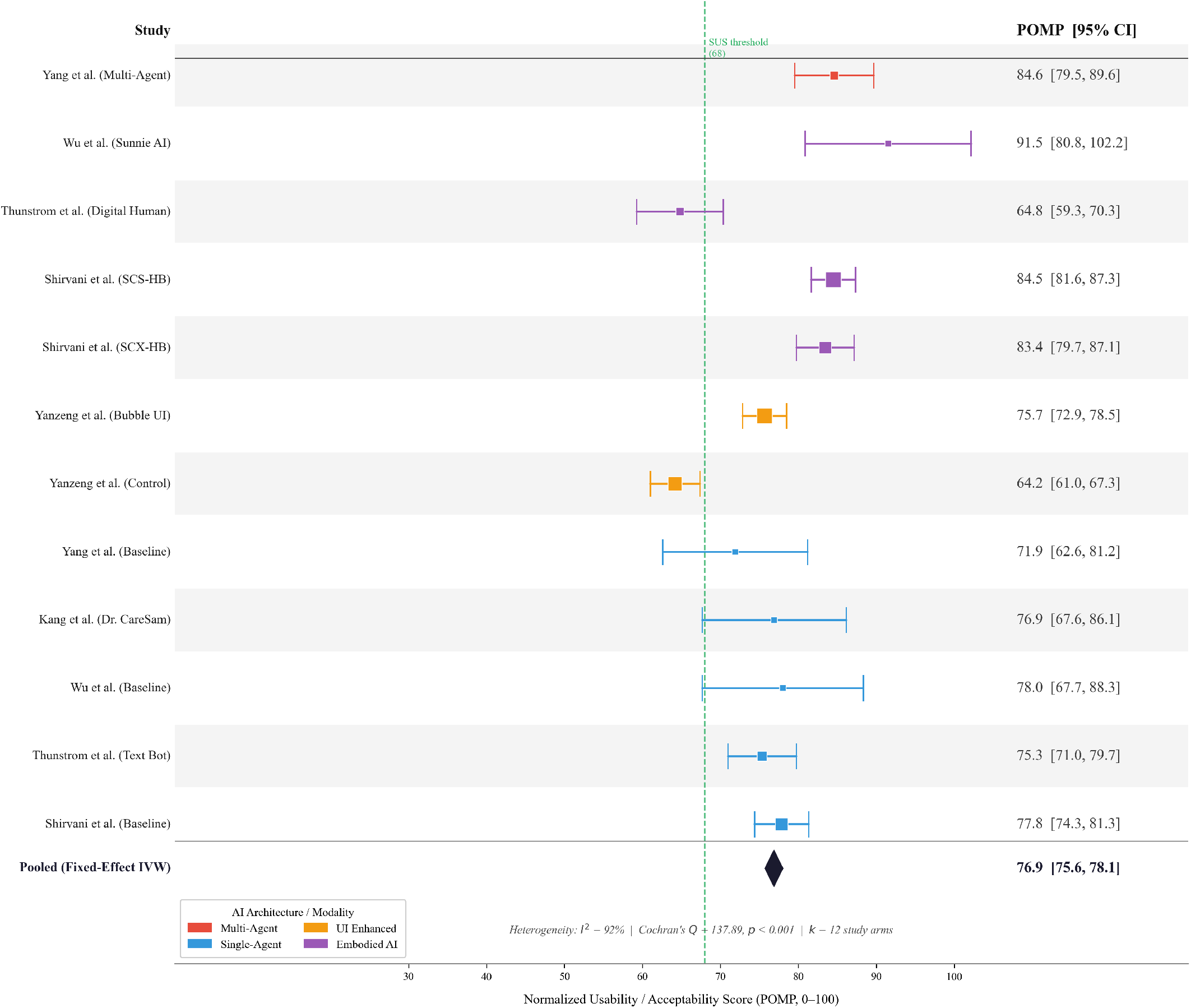
Forest plot of POMP-normalised usability and acceptability scores across 12 study arms from 6 included studies. Squares indicate point estimates with size proportional to inverse-variance weight; horizontal bars represent 95% confidence intervals. The dashed vertical line marks the SUS acceptability threshold (POMP = 68). The pooled fixed-effect estimate (diamond) is 76.9 (95% CI: [75.6, 78.1]); *I*^2^ = 92% reflects substantial heterogeneity driven primarily by architectural and modality differences across systems rather than baseline LLM capability.

Intra-study comparisons reveal that these design choices substantially moderate acceptability. Yang et al. [94] found that a multi-agent architecture (POMP = 84.6, 95% CI: 79.5–89.6) significantly outperformed a single-agent baseline (POMP = 71.9, 95% CI: 62.6–81.2), suggesting that distributing conversational roles across specialised agents enhances perceived quality of psychological support. Wu et al. [**?**] showed that endowing an LLM agent with anthropomorphic traits (POMP = 91.5) yielded markedly higher perceived utility than a non-anthropomorphic baseline (POMP = 78.0), consistent with the social-presence literature. Conversely, Thunström et al. [96] surfaced an *uncanny valley* effect: a text-only chatbot (POMP = 75.3) outperformed a fully embodied digital-human variant (POMP = 64.8), implying that imperfect photorealistic embodiment may undermine rather than enhance user trust. Taken together, these findings indicate that while LLM-driven agents as a class consistently clear the acceptability threshold, the front-end interaction paradigm—particularly the choice between multi-agent orchestration and embodied presentation—remains a critical determinant of sustained user engagement in digital mental health applications.

### Datasets and benchmarks for systematic comparison

Beyond evaluating individual systems in isolation, a growing subset of studies builds empirical datasets, annotation schemes and benchmark frameworks designed to compare model performance, target specific capabilities and track progress over time. MDD-5k, for instance, provides the first labeled dataset for benchmarking Chinese mental disorder diagnosis, combining synthetic diagnostic dialogues grounded in real, anonymized patient cases with structured labels mapped to DSM/ICD categories [80]. KMI offers the first synthetic dataset specifically designed to support chatbots grounded in motivational interviewing, generating 1,000 high-quality dialogues that reflect therapist decision policies as well as language-level realizations [93].

Building Trust in Mental Health Chatbots proposes a multi-dimensional evaluation framework that explicitly links safety, reliability and perceived expertise, pairing automated scenario-based tests with structured human ratings across trust-related constructs [167]. MindEval pushes this direction further by targeting multi-turn therapy conversations and formalizing a workflow that incorporates PhD-level licensed clinical psychologists into the annotation pipeline, creating a benchmark where LLM-generated therapeutic dialogues are scored along clinically meaningful dimensions [38]. These benchmark-oriented efforts move the field toward more standardized, comparable and clinically grounded evaluations, although they remain relatively few compared to task-specific studies.

### Artificial users and simulation-based evaluation

To preserve patient privacy and enable controlled testing, several studies introduce *artificial users* that emulate real patients’ behavior and reasoning based on clinical vignettes. In one such study, the authors vary depression severity, personality traits and attitudes to generate 48 simulated users interacting with an LLM-based chatbot; the resulting dialogues are then evaluated by ten professional psychotherapists using standardized rating scales for therapeutic quality, alliance and safety [168].

Another line of work simulates both doctor and patient agents in multi-agent setups. One study first assesses applicability using automated metrics such as recall, accuracy and task-specific ratios, and then introduces a human evaluation rubric for doctor agents along four dimensions: fluency, empathy, expertise and engagement. And for patient agents along two dimensions: resemblance to realistic patient behavior and rationality [169]. Similarly, Sibyl [170] evaluates empathetic dialogue generation via a layered strategy: it begins with automatic language benchmarks (for example, BLEU and ROUGE) and then asks professional annotators to apply a structured rubric to refine judgments of language ability, empathy and appropriateness. These simulation-based approaches allow systematic manipulation of user characteristics and scenario difficulty, but they must be carefully validated to ensure that artificial users accurately reflect real patient heterogeneity and communication styles.

### Safety- and reliability-centered evaluation trajectories

Beyond benchmarking a single system in isolation, a growing set of studies adopts a comparative angle and evaluates LLM-based agents across multiple tasks or dimensions. In these cross-task evaluations, safety is typically treated as the first-order concern, with suicidality and acute risk scenarios foregrounded. Several studies therefore design simulated suicidal-risk situations and explicitly test how chatbots or general-purpose LLMs respond when users express suicidal thoughts or behaviors, using controlled prompts to examine whether the models provide appropriate, non-harmful guidance and escalation cues [166], [171]. Building on this line of work, other frameworks broaden the safety scope from a single high-stakes scenario to a more comprehensive psychosocial risk profile. DialogGuard, for example, extends evaluation beyond suicidality by operationalizing five additional dimensions of harm: privacy violations, discriminatory behavior, mental manipulation, psychological harm and insulting behavior [34].

Once immediate harm risks are addressed, some studies shift the focus from “is it safe?” to “can it be trusted?”, emphasizing reliability and clinical soundness over prolonged interactions. Building Trust in Mental Health Chatbots curates a set of benchmark questions paired with ideal responses and compares model outputs against these references to assess consistency, correctness and trust-related properties [167]. MindEval targets multi-turn therapy conversations specifically, integrating structured clinical rubrics with automated scoring pipelines to evaluate how well LLMs sustain coherent, empathic and therapeutically appropriate dialogue over time [38]. Other evaluations examine specific modalities, such as voice-enabled generative AI for mental health support, assessing not only task performance but also conversational flow, perceived alliance and user comfort in spoken interactions [165].

Taken together, these efforts reflect a widening evaluation trajectory: from narrow offline metrics and generic language benchmarks, to scenario-based safety tests, especially around suicidality, to broader psychosocial harm frameworks and reliability-oriented measures grounded in clinical expertise. However, fully realizing the promise of clinician and patient-in-the-loop validation will require more prospective, longitudinal studies in real care settings, with clear reporting of cohorts, outcomes and failure modes. In later sections, we build on this trajectory to propose an evaluation ladder tailored to mental health agents, linking specific task claims to appropriate levels of evidence and governance requirements.

#### Standardizing the human loop

While the preceding subsections document a definitive shift toward human-in-the-loop validation, the field remains fragmented regarding the specific composition of the evaluation panel and the criteria for success. Table VII synthesizes protocols from five representative systems, delineating three distinct methodological paradigms based on the system’s intended utility. For high-stakes diagnostic tasks, systems such as MDD-5k and MentalRAG rely on **Expert-Led Clinical Audits**, recruiting licensed clinicians to rigorously assess safety and guideline adherence, thereby prioritizing clinical validity over user satisfaction. In contrast, wellness-focused agents like GymBuddy and Nova favor **Outcome-Oriented Field Trials**, deploying longitudinal RCTs to measure tangible behavioral changes (e.g., exercise adherence) alongside self-reported engagement. Occupying a third niche are simulation platforms like SocialSim, which utilize **Scale-Driven User Studies** to leverage crowdsourced ratings, effectively trading clinical depth for the statistical power needed to assess conversational realism. This methodological diversity reflects a broader spectrum ranging from the strict control of small-*N* expert review to the ecological validity of naturalistic deployment, suggesting that the optimal protocol is dictated by the agent’s specific risk profile rather than a single universal standard.

**TABLE VII.** Taxonomy of Human-in-the-Loop Evaluation Protocols.

| System | Evaluator profile (*Who*) | Study design protocol (*How*) | Evaluation criteria (*What*) |
| --- | --- | --- | --- |
| **MDD-5k** [80] | 3 licensed psychiatrists (5 years clinical experience) | Blind review of 100 synthetic diagnostic dialogues; inter-rater reliability (Fleiss' $\kappa$) computed | Diagnostic accuracy (DSM-5 alignment); symptom coverage; clinical plausibility; safety (harm detection) |
| **KMI** [93] | 2 MI-certified therapists and 5 clinical psychology PhD students | Pairwise comparison of 50 MI dialogues (human – synthetic); Likert scales (1–5) on 6 dimensions | MI adherence (MITI codes); empathy; open-ended questioning; reflective listening; goal elicitation; naturalness |
| **GymBuddy** [172] | 124 university students (randomized controlled trial; 8-week intervention) | Pre-post surveys (PHQ-9; exercise frequency); weekly engagement logs; exit interviews ($n = 20$) | Mood improvement ($\Delta$ PHQ-9); exercise adherence (days per week); perceived usefulness; trust; dropout rate |
| **Nova** [173] | 89 older adults (age $\geq 65$; quasi-experimental; 12-week deployment) | Baseline and endpoint assessments (loneliness scale; SUS); usage analytics (session count; duration); semi-structured interviews ($n = 15$) | Loneliness reduction; usability (SUS score); engagement (sessions per week); perceived companionship; safety incidents (self-reported) |
| **SocialSim** [20] | 500 crowdworkers (Amazon Mechanical Turk; diverse demographics) | Each rater evaluates 10 multi-turn conversations (agent – human baseline); 5-point Likert on 4 dimensions; attention checks | Conversational realism; empathy; coherence; appropriateness; preference (agent – human) |

**Note:** **MI:** Motivational Interviewing; **MITI:** Motivational Interviewing Treatment Integrity; **PHQ-9:** Patient Health Questionnaire (9-item depression scale); **SUS:** System Usability Scale.

## Translational risks, ethics, and governance: aligning agentic design with accountability and regulation

Risk in mental health AI is often treated as a monolithic concept. To support precise governance, we decompose the probability of clinical harm Pr(Harm) into a chain of conditional failures—the “Swiss Cheese Model” of AI safety. Given an input *x*, a potential unsafe response *a*_unsafe_, and a user context 𝒞_*user*_, the total risk is the product of generation, detection failure, and compliance:

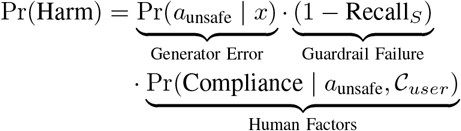

Here, Recall_*S*_ represents the sensitivity of the safety overlays (such as the specific *ϵ*-thresholds defined in the MAS architecture) in identifying toxic outputs. This factorization clarifies that reducing harm requires a **layered defense strategy**: (i) minimizing the generation of unsafe advice via alignment; (ii) maximizing Recall_*S*_ through dedicated critic agents; and (iii) simultaneously lowering the conditional probability of user compliance through interface friction, disclaimers, or human-in-the-loop escalation protocols.

Our audit reveals that translational risks in mental health agents arise from both model-level errors and how systems are embedded into workflows, presented to users and governed over time. We group key concerns into five broad categories: (i) hallucination and unsafe advice, (ii) bias amplification and inequity, (iii) privacy and secondary data use, (iv) crisis escalation and risk management, and (v) scope creep from wellness support into de facto diagnosis or treatment. Agentic design choices such as tool use, long-term memory, multiagent supervision and escalation logic, interact with each of these risk categories and therefore must be explicitly linked to accountability mechanisms and regulatory pathways. First, hallucination and unsafe advice remain central hazards, especially when agents are used in high-stakes contexts such as suicidality, acute crises or medication management. Systematic evaluations of LLM safety in addressing depression and suicidal ideation have shown that even clinically branded chatbots frequently omit crisis resources, provide generic or ambiguous guidance and occasionally produce content that could be interpreted as dismissive or invalidating [125]. Cross-model comparisons of general-purpose LLMs and domain-specific systems likewise find substantial variability in safety performance, with some models offering appropriate deescalation and others failing to recognize risk cues in user messages [166], [171]. Domain-specific safety frameworks such as DialogGuard and PsySafe attempt to mitigate these risks by explicitly modeling psychosocial harms, for example, privacy violations, discriminatory behavior, psychological harm, and stress-testing agents in multi-agent simulations [34], [76]. However, most production systems still lack continuously monitored safety layers, and few report post-deployment incident data.

Second, bias amplification and inequity arise when models are trained on demographically skewed data or when their interaction styles reflect majority cultural norms. As noted earlier, many systems are developed and evaluated primarily in North American, European or East Asian contexts and often on convenience samples of young, digitally literate users. Without explicit auditing across age, sex, gender identity, race/ethnicity and culture, agents may systematically underperform or behave in culturally insensitive ways for underrepresented groups. Recent work on cultural prompting and cross-cultural empathy demonstrates that default LLM behavior often aligns more closely with Western conversational norms, and that targeted adaptation is required to improve perceived empathy for users from non-Western backgrounds [153]. From a governance perspective, this implies that fairness audits must be part of the core evaluation plan rather than an optional add-on, and that deployment claims should be explicitly limited to populations for which adequate evidence exists.

Third, privacy and secondary data use are persistent concerns in mental health contexts, where conversations often include highly sensitive information about trauma, self-harm, relationships and illegal behavior. Many systems address privacy primarily through dataset-level anonymization and de-identification [80], [157], but a growing subset treat privacy as a first-class design objective. MentalRAG, for example, stores user data exclusively on local devices, protects it with encryption and uses on-device processing so that sensitive information never leaves the user’s environment [82]. Other systems integrate privacy-preserving retrieval modules that anonymize personal identifiers before constructing context for RAG pipelines, and only re-inject identifiers at the final step to preserve personalization [71]. While these approaches reduce some risks of data leakage, they do not fully address issues of secondary use, for example, using logs to train future models, or cross-border data transfer. Transparent data lineage, explicit consent for secondary use and user-level controls, for example, “right to forget” options, remain uncommon in current deployments.

Fourth, crisis escalation and risk management are inadequately addressed in many evaluated systems. In one study, among 25 chat-based agents evaluated using a standardized PHQ-9 depression screener, only 2 provided crisis resources, such as suicide hotlines, to simulated users who screened positive for severe depression [125]. This finding, echoed in other safety audits of mental health chatbots, underscores substantial gaps in safety-oriented behavior even when models correctly recognize high-risk responses. Effective governance therefore requires explicit, tested escalation protocols: for example, defining when and how agents should (i) provide crisis hotline information, (ii) encourage immediate contact with emergency services, (iii) refuse to continue without a human in the loop or (iv) notify a supervising clinician in integrated care settings. Multi-agent architectures can support this by separating “therapeutic” agents from “safety critic” or “risk sentinel” agents that continuously monitor conversations and can override or block unsafe responses [76], [174]. However, these safety agents must themselves be validated against clinical standards and regulatory expectations.

Finally, scope creep and misuse raise subtler but equally important translational risks. As models become more capable and agentic, there is a growing gap between how developers intend systems to be used, for example, as wellness companions or decision-support tools, and how users may actually deploy them, for example, as de facto diagnosticians, sources of legal advice or tools for gaming assessments. One line of research has already shown that students can leverage ChatGPT to feign ADHD symptoms in self-report instruments, potentially distorting educational accommodations and clinical assessments [114]. Related concerns have been raised about malingering in forensic or legal contexts, where individuals might use AI to simulate or exaggerate psychiatric symptoms to avoid legal consequences. These risks motivate clearer scoping statements, user-facing warnings and technical safeguards, for example, limiting certain outputs, detecting repeated “test-taking” patterns, or requiring clinician verification for high-stakes decisions.

From a regulatory standpoint, these risk categories intersect with evolving frameworks for medical devices and AI governance. When agents make diagnostic or treatment recommendations, or when their outputs are integrated into clinical records, they are likely to fall under medical device regulations and require evidence of safety, effectiveness, and post-market surveillance. Even when positioned as “wellness” tools, systems that influence risk behaviors—for example, suicide and substance use—may warrant additional oversight.

Our review suggests that aligning agentic design with accountability to structurally minimize Pr(Harm) requires: (i) explicit mapping of agent roles to responsibility, for example specifying which agent plans and which agent audits safety (thereby maximizing the guardrail sensitivity Recall_*S*_) and where escalation to human clinicians is mandated; (ii) documentation of data provenance, limitations, and intended use to bound generator uncertainty; (iii) ongoing monitoring with mechanisms for incident reporting, model updates, and user recourse to detect shifts in user vulnerability (𝒞_*user*_); and (iv) conducting multiple rounds of real-world testing before releasing systems to the public. Without such governance structures, the field risks repeating historical patterns in digital mental health, where promising tools are deployed widely before their risks and benefits are fully understood.

## Toward clinically accountable, role-aware multi-agent mental health AI

Throughout this Review, we have emphasized that the label *agent* is used inconsistently across the literature: some works describe any conversational interface as an “agent”, while others reserve the term for systems with explicit planning, tool use, memory and multi-agent coordination. For clinicians and regulators, these distinctions are not merely semantic. Agentic features such as autonomous goal selection, tool calling and inter-agent debate, change both what a system can do and how responsibility should be assigned when things go wrong.

By auditing architectures, data provenance, demographics, task claims and evaluation rigor within one unified framework, we aim to provide a common map that allows researchers, clinicians and regulators to judge readiness across diverse mental health agents.

Several cross-cutting themes emerge from this synthesis. First, current applications are heavily concentrated on depression and anxiety, with relatively sparse coverage of severe mental illness, neurodevelopmental disorders, substance use, sleep–wake disorders and complex comorbidity. This topical skew reflects data availability and perceived risk, but it also creates blind spots: agents trained and evaluated primarily on mild-to-moderate mood and anxiety conditions may not generalize safely to individuals with psychosis, bipolar disorder, cognitive impairment or multi-morbidity. Second, many systems exhibit a mismatch between their technical sophistication, for example, complex multi-agent workflows, multimodal sensing, and the modesty of their evaluation, which often relies on offline metrics, short-term user satisfaction surveys or vignette-based safety tests. Third, demographic and cultural coverage is uneven, with evidence concentrated in a small number of regions and populations, limiting claims about global applicability and fairness.

Against this backdrop, our review points toward a design paradigm centered on *role-aware* multi-agent systems with transparent retrieval, explicit personalization, embedded risk assessment and routine clinician oversight. In such systems, agents are assigned well-defined roles such as history-taking, psychoeducation, safety monitoring, evidence retrieval or documentation drafting, and coordination mechanisms make these roles explicit in both system logs and user interfaces. Retrieval components should expose their sources and limitations, so that clinicians and patients can see which guidelines or datasets underpin specific recommendations. Personalization modules should operate within clearly defined consent and privacy boundaries, avoiding opaque long-term profiling. Safety critics and risk sentinels should be architecturally separated from therapeutic agents and empowered to override or block unsafe actions. Most importantly, clinician oversight should be built into the workflow for high-stakes tasks, so that human experts remain ultimately accountable for diagnostic and treatment decisions.

Achieving clinically accountable, role-aware mental health AI also requires a shift in evaluation culture. Instead of treating offline benchmarks as sufficient evidence, multi-agent systems should be assessed under realistic, longitudinal conditions with both clinician and patient participation. This includes measuring not only performance on narrowly defined tasks, but also downstream outcomes such as care engagement, treatment adherence, therapeutic alliance, symptom trajectories and unintended harms. Evaluation frameworks like MindEval, Building Trust in Mental Health Chatbots and PsySafe [38], [76], [167] illustrate how structured rubrics, synthetic users and expert annotators can support more clinically meaningful assessments. Extending these efforts into prospective clinical trials and routine-care audits is a critical next step.

### Outlook

Looking ahead, we anticipate that next-generation mental health agentic systems will evolve from monolithic chat interfaces into supervised, role-aware multi-agent collaborators that operate within, and not outside, clinical pathways. Rather than positioning a single LLM as an all-purpose counselor, future systems are likely to comprise ensembles of specialized agents for tasks such as triage, psychoeducation, safety monitoring, guideline retrieval, documentation drafting and patient coaching. These agents will need to interoperate with electronic health records, appointment and referral systems and other components of real-world care infrastructure, with clear boundaries around which actions can be taken autonomously and which require clinician sign-off.

Several priority directions emerge from our framework. First, *multimodal evidence integration with verifiable provenance* will be crucial. High-quality systems should be able to combine text, speech, facial expressions, physiological signals and structured clinical data, while documenting where each signal comes from, how it is processed and what uncertainties remain. While proposing these systems, some studies may face data scarcity. Multimodal mental health data are often collected in small, non-diverse cohorts due to data sharing and privacy constraints, whereas large models typically benefit from substantial and diverse training data. Federated learning (FL) [175], [176] and related aggregation approaches have therefore emerged as a potential solution, enabling systems to benefit from more diverse and abundant data without direct data sharing.

Second, *demographic and cultural broadening* is essential. Future work should explicitly recruit adolescents, older adults and underrepresented communities, and should include cross-cultural validation of both language behavior and clinical outcomes. Culturally adapted frameworks, such as those developed for Persian or for non-Western clinical settings [151], [154], offer promising starting points but remain the exception rather than the norm.

Third, *embedded safety critics and policy supervisors* should become standard rather than experimental features. Domain-specific constitutional AI approaches and multi-agent safety frameworks like DialogGuard and PsySafe [34], [76], [174] point toward architectures where safety policies are explicit, inspectable and enforceable at run time. Such policies will need to be co-designed with clinicians, ethicists, legal experts and patient advocates, and kept up to date as regulations and clinical guidelines evolve. Fourth, *prospective, real-world studies* are needed to measure both benefit and harm in routine care, including unintended consequences such as over-reliance on AI, deskilling, inequitable access or changes in help-seeking behavior. These studies should incorporate pre-registered protocols, independent oversight and transparent reporting of negative as well as positive findings.

Finally, shared benchmarks and governance frameworks should more closely reflect the realities of clinical practice. Benchmarks ought to specify clinical tasks, require transparent data lineage and incorporate clinician and patient participation in annotation, scoring and interpretation. Governance frame-works should align technical documentation, such as model cards, data statements and safety reports, with regulatory requirements for medical devices and AI systems, including post-deployment monitoring and incident response. If these steps are taken, the gap between physician and AI perspectives can narrow, enabling agentic mental health AI to move from experimental prototypes toward responsible, clinically integrated tools that support—not supplant—human care.

## Data Availability

All data produced are available online at https://github.com/PAI-CUHK/Mental-Health-Agents-Survey

## Acknowledgments

This work was supported by the Research Grants Council Theme-based Research Scheme (RGC-TRF ref. no. T14-412/25-R), the CUHK Vice-Chancellor Early Career Professorship Scheme (ref. no. 441), the CUHK Teaching Development and Language Enhancement Grant 2025-28 (TDLEG ref. no. ATD100), National Natural Science Foundation of China (ref. no. 62573271), and Major Basic Research Project of Shandong Provincial Natural Science Foundation (ref. no. ZR2025ZD28).

## Author contributions

**L.Z**.: Writing - review & editing, Writing - original draft, Validation, Software, Resources, Methodology, Investigation, Formal analysis, Data curation. **W.W**.: Writing - review & editing, Writing - original draft, Validation, Software, Resources, Methodology, Investigation. **W.T**.: Writing - review & editing, Writing - original draft, Data Curation. **B.C**.: Writing - review & editing, Writing - original draft, Data Curation. **X.LIN**: Writing - review & editing, Writing - original draft, Data Curation. **Z.W**.: Writing - review & editing, Writing - original draft, Data Curation. **H.Y**.: Writing - review & editing, Writing - original draft, Data Curation. **X.LI**: Writing - review & editing, Writing - original draft, Data Curation. **J.J**.: Writing - review & editing, Writing - original draft. **S.H**.: Writing - review & editing, Writing - original draft. **Y.ZHONG**: Writing - review & editing, Writing - original draft. **Y.ZHENG**: Writing - review & editing, Writing - original draft. **J.S**.: Writing - review & editing, Writing - original draft. **A.H**.: Writing - review & editing, Writing - original draft. **L.L**.: Writing - review & editing, Writing - original draft, Data Curation. **J.Z**.: Writing - review & editing, Writing - original draft, Data Curation. **W.H**.: Writing - review & editing, Writing - original draft, Data Curation. **N.Y.C**.: Writing -review & editing, Writing - original draft, Resources, Supervision, Funding acquisition. **L.L**.: Writing - review & editing, Writing - original draft, Validation, Supervision, Resources, Methodology, Investigation. **Y.K.W**.: Writing - review & editing, Writing - original draft, Validation, Supervision, Resources, Methodology, Investigation, Funding acquisition. **X.M**.: Writing - review & editing, Writing - original draft, Validation, Supervision, Resources, Project administration, Methodology, Investigation, Data curation, Formal analysis, Conceptualization, Funding acquisition. **L.F**.: Writing - review & editing, Writing - original draft, Validation, Supervision, Resources, Project administration, Methodology, Investigation, Data curation, Formal analysis, Conceptualization, Funding acquisition.

## Supplementary Information

### Overview

This appendix describes the literature search and screening process, annotation framework, inter-rater agreement procedure, and the review’s positioning relative to existing surveys. The aim is to make the review workflow transparent and reproducible without altering the substantive criteria or coding content used in the main study.

### Data Collection and Literature Selection

*Search scope and eligibility criteria:* We conducted comprehensive searches across arXiv, medRxiv, OpenAlex, OSF Registries, PubMed/MEDLINE, Scopus, SpringerLink, and Web of Science Core Collection, covering the period from January 1, 2023 to December 31, 2025. Records were considered eligible if the title, abstract, or keywords simultaneously referenced three concept blocks: (1) an interactive system or interface, (2) an AI or model family term, and (3) a mental-health domain term.

*Rationale for the three-term eligibility design*. The three-block requirement shown in Table VIII was applied to ensure that retrieved records concern AI-enabled agentic systems situated in mental-health contexts, including both conversational and non-conversational agents, while limiting concept drift toward non-agent interventions or non-AI digital tools.

**TABLE VIII.** Rationale for requiring three concurrent concept blocks in title, abstract, or keywords.

| Concept block | Anchor terms used in query | What this block is meant to capture | Why we anchor and what we avoid |
| --- | --- | --- | --- |
| Interactive system / interface | *agent OR chatbot* | Agentic AI applications: systems described as an agent or chatbot that take an autonomous or semi-autonomous task role in a specific workflow. | **Anchor for cross-source reproducibility:** “chatbot” and, during 2023–2025, “agent” are the most consistently used metadata descriptors across clinical and computer-science venues. **Avoid:** enumerating heterogeneous subtype labels such as conversational agent, dialogue system, and virtual therapist, which are inconsistently indexed and are often absent from title, abstract, or keywords. |
| AI / model family | *AI OR LLM OR large language model* | Records explicitly involving AI systems. We included LLM / large language model specifically because some papers refer to LLMs without using the broader “AI” label. | **Anchor for technical specificity:** requiring an AI/LLM descriptor ensures that retrieved records involve an actual computational AI component. **Avoid:** general digital mental-health tools that may be described as “chatbots,” “apps,” or “interventions” without any AI implementation. |
| Mental-health domain | *mental health OR psychiatry OR psychology* | Records explicitly situated in mental-health contexts, including the evaluation, application, or discussion of AI or LLM systems for mental-health-related purposes, rather than general wellness coaching or unrelated conversational AI. | **Anchor for domain validity:** requiring a mental-health descriptor ensures that retrieved records are substantively connected to mental health as a clinical or psychological domain. **Avoid:** concept drift toward general health or wellbeing applications, omission caused by narrow diagnosis-specific keywords, and inflated false positives from broad health-related terms. |

The Boolean structure of the search keywords was:

~~~
(Agent OR Chatbot) AND (AI OR
LLM OR Large Language Model) AND
(Mental Health OR Psychiatry OR
Psychology)
~~~

#### Deduplication procedure

To remove duplicates across databases, we applied a two-stage deduplication procedure. First, we normalized bibliographic strings by lowercasing and trimming whitespace and punctuation, and we constructed a comparison key by concatenating each record’s title and author list into a single string. Second, we computed the Jaccard similarity between pairs of records based on token sets derived from these concatenated strings.

Concretely, for two records *A* and *B*, let *T* (*A*) and *T* (*B*) denote the sets of tokens extracted from the concatenated *title+authors* strings. The Jaccard similarity was computed as

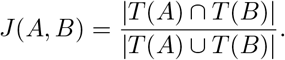

Any pair with *J*(*A, B*) *>* 0.8 was flagged as a potential duplicate. All flagged pairs were then manually inspected by a single reviewer to confirm duplication. For confirmed duplicates, we retained the earliest appeared record and removed redundant entries. When there was ambiguity, the record was retained for downstream screening to avoid erroneous exclusion.

#### PRISMA flow diagram

The PRISMA flow diagram in Figure 12 illustrates the number of studies retained and excluded at each stage of the screening process.

**Fig. 12.**
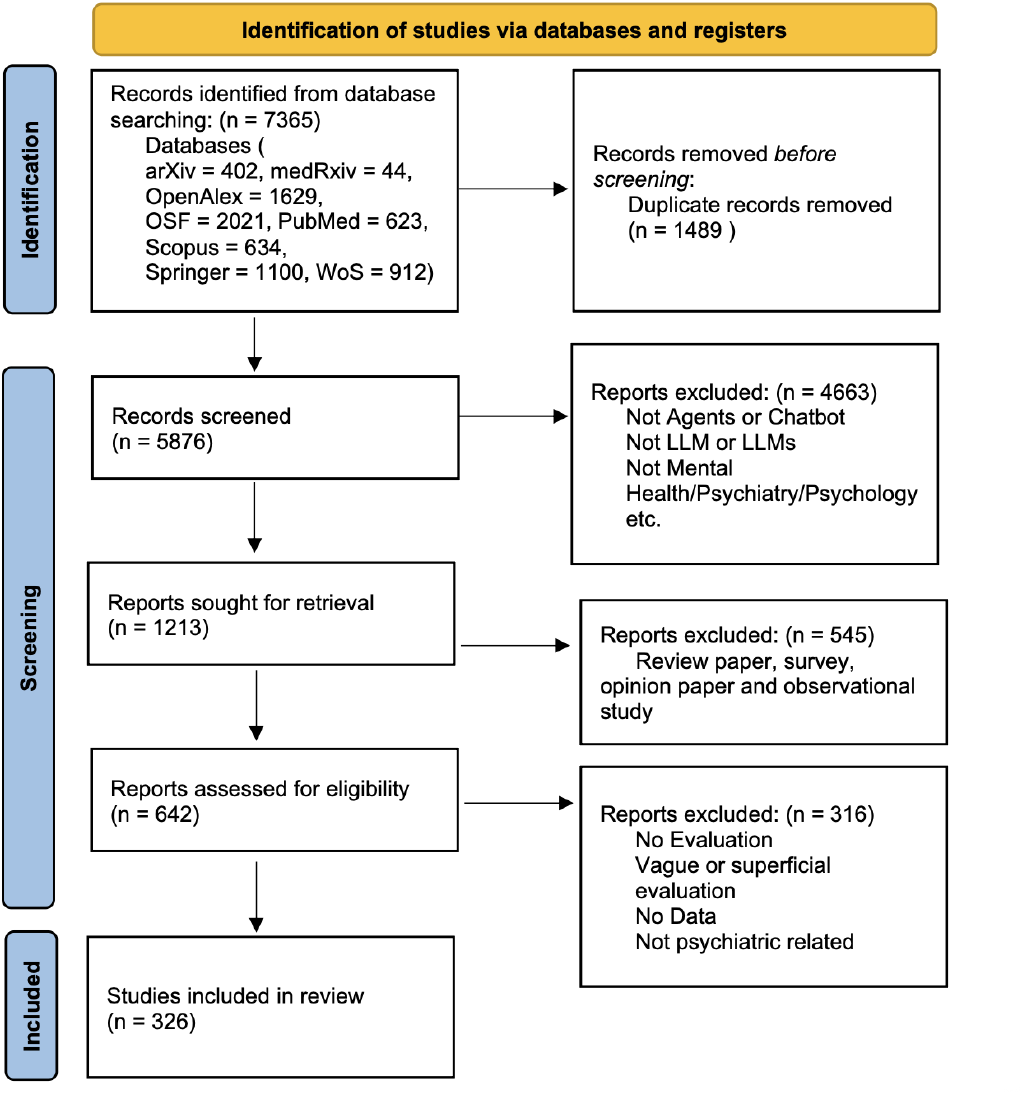
PRISMA flow diagram.

### Annotation Framework

After screening, 326 articles remained for full review. Each article was coded across six dimensions: system type, data scope, mental health focus, demographics, downstream tasks, and evaluation type. Unless otherwise specified, coding was multi-label, meaning that more than one label could be assigned when applicable.

#### 1. System Type

System type captured how the system was built and how it operated. This dimension included **(a) base model type**, including **Rule-based models, Machine learning models, Deep learning models, Small pre-trained model**, and **Foundation model**; **(b) interface type**, including **Chatbot** and **Non-chat agent**; **(c) workflow composition**, including **Non-agent, Single agent**, and **Multi-agent**; **(d) multi-agent architecture** [68], including **Flat architecture, Hierarchical architecture, Team architecture**, and **Hybrid architecture**; and **(e) agentic role**, including **Retrieval, Personalization, Risk assessment, Tool use**, and **Other**. Here, interface type distinguishes conversational from non-conversational systems, workflow composition distinguishes the degree of agent autonomy and coordination, and agentic role captures the main operational function of the agent within the system.

#### 2. Data Scope

Data scope captured the type and origin of the data used by each study. This dimension included **(a) modality**, including **Text, Audio, Images**, and **Video**; and **(b) source of data**, including **Clinically collected data, Electronic health records (EHRs), Lab tests, Elicited evidence, Interview / questionnaire, Interactional / conversational**, and **Generated / synthetic**. Within elicited evidence, we further noted study designs such as **Systematic reviews / meta-analyses, Randomized controlled trial (RCT), Cohort study, Case series and reports**, and **Cross-sectional / prevalence study**. In this framework, modality refers to the format of the data, whereas source of data refers to how the data were generated or collected.

#### 3. Mental Health Focus with ICD-11 Groupings

Mental health focus captured the diagnostic or disorder domain addressed in each paper, using ICD-11 groupings [107]. This dimension included **06 Mental, behavioural or neurodevelopmental disorders**, covering **6A0 Neurodevelopmental disorders, 6A2 Schizophrenia or other primary psychotic disorders, 6A4 Catatonia, Mood disorders, 6B0 Anxiety or fear-related disorders, 6B2 Obsessive-compulsive or related disorders, 6B4 Disorders specifically associated with stress, 6B6 Dissociative disorders, 6B8 Feeding or eating disorders, 6C0 Elimination disorders, 6C2 Disorders of bodily distress or bodily experience, Disorders due to substance use or addictive behaviours, 6C7 Impulse control disorders, 6C9 Disruptive behaviour or dissocial disorders, 6D1 Personality disorders and related traits, 6D3 Paraphilic disorders, 6D5 Factitious disorders, 6D7 Neurocognitive disorders**, and **6E2 Mental or behavioural disorders associated with pregnancy, childbirth or the puerperium**. ICD-11 groupings were used to standardize disorder coding across heterogeneous terminology in the included studies.

#### 4. Demographics

Demographics captured the population characteristics reported in the included studies. This dimension included **(a) age group**, including **Childhood, Adolescence, Young adulthood, Middle adulthood, Old**, and **Age unspecified**; **(b) study location**, including **Country / Nation**; and **(c) sex**, including **Male, Female**, and **Unspecified**. These labels were used to summarize the populations represented across studies and to identify gaps in demographic reporting.

#### 5. Downstream Tasks

Downstream tasks captured the main intended application of the system. This dimension included **Screening / triage, Clinical decision support, Therapeutic interventions, Documentation generation, Ethical / legal support, Education & simulation**, and **Other**. These categories distinguish whether the system was intended for assessment, treatment, documentation, professional support, or educational use.

#### 6. Evaluation Type

Evaluation type captured how system performance or usefulness was assessed. This dimension included **Automated evaluation**, with subtypes **Applicability evaluation** and **Language evaluation**; **Human evaluation**, with subtypes **Physician / expert involvement** and **Patient / user involvement**; and **Evidence & benchmarking**. Here, automated evaluation refers to metric-based assessment, human evaluation refers to judgment by experts or end users, and evidence & benchmarking refers to benchmark construction or evaluation-framework development.

### Inter-reviewer Agreement

For reliability, we randomly sampled 15 papers and asked three reviewers to independently annotate each article, using the model’s suggestions only as optional hints. Across 435 tag assignments, Krippendorff’s alpha reached approximately 0.81, indicating strong agreement. We computed Krippendorff’s alpha as

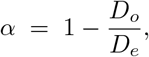

where *D*_*o*_ is the observed disagreement and *D*_*e*_ is the disagreement expected by chance. The observed disagreement is

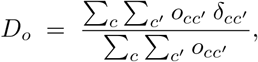

and the expected disagreement is

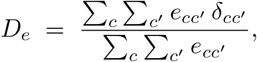

where 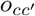 and 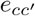 denote the observed and expected coincidence counts for category pairs (*c, c*^*′*^), and 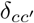 is the distance between categories, with 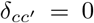 if *c* = *c*^*′*^ and 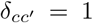 otherwise for nominal labels. Given this agreement level, we proceeded with single-reviewer annotation for the remaining papers, while escalating ambiguous cases for group discussion and adjudication.

### Comparison with Existing Reviews and Surveys

Several recent reviews have examined the use of AI, chatbots, or large language models in mental health and psychiatry, but they differ from the present review in scope, granularity, and analytic framework.

Hua et al. [177] reviewed the evolution of mental-health chatbots from rule-based systems to LLM-based systems, with an emphasis on conversational support, psychoeducation, symptom checking, and staged maturity from early feasibility to implementation. Their review provides a useful technology lineage and highlights safety and regulatory considerations, but it remains focused primarily on chatbot systems and does not formalize planner-based behavior, tool use, or multiagent workflow composition. In contrast, our review extends beyond chatbots to include broader mental-health *agents*, including non-chat and background systems, and explicitly codes workflow composition and agentic roles such as retrieval, personalization, risk assessment, and tool use.

Guo et al. [178] surveyed large language model applications in mental health, covering topics such as detection, triage, suicide risk, and mental-health advice. Their review discusses LLMs mainly in terms of task categories and use cases, and it notes important concerns such as hallucination, safety, and the limited number of prospective studies. However, it does not provide a structured taxonomy of data provenance or workflow structure. By comparison, our review introduces a formal *Data Scope* framework, including clinically collected data, electronic health records, laboratory data, elicited self-report, interactional data, and synthetic data, while also distinguishing non-agent, single-agent, and multi-agent workflows.

Omar et al. [179] focused on applications of large language models in psychiatry, particularly diagnostic support, suicide risk, and clinician education. Their discussion is largely organized around narrative scenarios and assistant-style uses of GPT-like systems, with strong attention to bias, hallucination, responsibility, and medico-legal concerns. However, disorder areas, demographic coverage, and data types are not systematically coded. Our review differs by applying a reproducible taxonomy across studies, including ICD-11 mental-health groupings, demographic dimensions, downstream task categories, and evaluation types.

Na et al. [180] surveyed large language models in psychotherapy, organizing the literature around therapy stages such as assessment, diagnosis, and treatment. Their review is especially valuable for psychotherapy-oriented systems and highlights the need for longitudinal and theory-grounded evaluation. Nevertheless, it is centered primarily on therapeutic chatbot settings and does not generalize to broader agentic or tool-integrated ecosystems. Our review instead spans a wider set of mental-health applications, including triage, clinical decision support, documentation, simulation and education, and benchmarking, while also coding architecture, data provenance, and evaluation practice.

Chiu et al. [181] proposed a computational framework for evaluating LLM therapists against human therapy styles, focusing on structured behavioral dimensions such as motivational interviewing fidelity. This work contributes an important evaluation perspective for single therapeutic chatbot systems, using scripted interactional settings and clinician-informed criteria. However, it is not designed as a general review framework across system architectures or study types. Our review generalizes evaluation coding across diverse mental-health AI systems by distinguishing automated evaluation, applicability evaluation, language evaluation, and human evaluation involving experts or end users.

Kuhlmeier et al. [182] examined the safety and quality of an LLM-based behavioral activation chatbot for depression using a hybrid framework that combined simulated artificial users with psychotherapist assessment. This study is notable for its semi-synthetic interactional design and expert review, but it remains focused on a single chatbot setting and a narrow clinical domain. In contrast, our review systematically differentiates synthetic, interactional, and clinically derived data sources and classifies evaluation designs across a much broader range of mental-health agent systems.

Taken together, prior reviews have made important contributions by characterizing chatbot evolution, summarizing LLM use cases, examining psychotherapy applications, or proposing targeted evaluation frameworks. However, they generally do not jointly audit system architecture, data provenance, demographics, disorder coding, and evaluation practice within a single framework. The present review addresses this gap by providing an integrated taxonomy of mental-health AI systems across three levels: *system design* (base model, interface, workflow, and agent roles), *data and population coverage* (modality, source, ICD-11 focus, age, sex, and geography), and *evaluation practice* (automated, human, and benchmarking-oriented assessment). In this sense, our review is positioned not only as a survey of applications, but also as a structured audit of clinical readiness in emerging mental-health agent systems.

## Notes

### Competing Interest Statement

The authors have declared no competing interest.

### Clinical Protocols

https://github.com/PAI-CUHK/Mental-Health-Agents-Survey

### Summary of Updates

This version of the manuscript has been revised to correct and update the institutional affiliations for authors Jie Sun, Yun Kwok Wing, Jiyuan Jiao, and Lizhou Fan.

